# Sex differences in the associations of genetic, sociodemographic and cardiovascular risk factors with depression in the Canadian Longitudinal Study on Aging (CLSA)

**DOI:** 10.1101/2023.04.10.23288267

**Authors:** Emilie Théberge, Jessica Dennis

**Affiliations:** University of British Columbia, Department of Medical Genetics

**Keywords:** Sex differences, major depressive disorder, cardiovascular disease, polygenic risk scores, CLSA

## Abstract

Major depressive disorder (MDD) is a leading cause of morbidity and disability worldwide, with approximately twice as many women reported to have a lifetime occurrence of MDD than men. MDD is a polygenic trait, wherein hundreds to thousands of common genetic variants with small effect sizes contribute to risk of disease. This study investigated sex differences in the risk factor comorbidity and genetic architecture of MDD in over 16,000 people aged 45-85 from the Canadian Longitudinal Study on Aging (CLSA), with 21% of females (n=1,741) and 12% of males (n=1,055) coded with MDD. Polygenic risk scores (PRS) for individuals were made using sex-stratified and non-sex-specific (“both-sexes”) UK Biobank genome-wide association study summary statistics data. Odds of MDD for the sex-specific PRSs, socioeconomic, lifestyle and clinical risk factors associated with cardiovascular disease risk were assessed using a multivariable logistic regression model for each sex. Significant sex-specific risk factor associations with odds of MDD were found in females (history of ischemic heart disease (OR 1.52 (1.14-2.01), hypothyroidism (OR 1.42 (1.25-1.63), not being partnered (OR 1.34 (1.17-1.52)), having diabetes (OR 1.30 (1.11-1.52)), and higher female sex-specific autosomal PRS (OR 1.10 (1.04-1.16))) and males (high blood pressure, OR 1.35 (1.04-1.47)). Significant differences were observed in the proportion of variables that contributed to the most to each model, evaluated by relative pseudo-R^2^ values. Age contributed the most to the model for both sexes (46.9% for females, 32.5% for males), wherein younger age was associated with higher odds of MDD. These results underscore the relevance for sex-disaggregating analyses of complex traits, like MDD, and the incorporation of clinical variables into models of MDD, in applications such as early detection and primary prevention.

## Introduction

Major Depressive Disorder (MDD) is a leading cause of morbidity and disability worldwide. An estimated 322 million people are affected by MDD annually, accounting for approximately 50 million years lived with disability (World Health Organization, 2017). A recent survey by Statistics Canada indicated that one in four (25%) Canadians aged 18 or over screened positive for depression, anxiety, or posttraumatic stress disorder, up from pre-pandemic estimates of population prevalence of 20% (Statistics Canada, 2021).

Mental illness does not exist in isolation from the body’s other systems. When the brain perceives stress, the hypothalamic-pituitary-adrenal (HPA) axis activates, and a cascade of biological processes ensues, affecting everything from digestion to cardiovascular output in a “fight-or-flight” response. In 2021, the American Heart Association provided a scientific statement on the “mind-heart-body” interconnections, providing an up-to-date overview of research describing how negative psychological health has causal influences on cardiovascular health and associated comorbidities like diabetes, dyslipidemia, hypertension, and obesity (Levine et al., 2021). The theory of “immunometabolic depression” describes the common mechanisms by which dysregulated homeostatic pathways in depressed patients can manifest as cardiometabolic traits, such as the effects of chronic low-grade inflammation and disrupted energy-regulating neuroendocrine signalling (Milaneschi et al., 2020).

MDD is a complex trait, whereby pathophysiology is influenced by both genetics and environment. Family and twin studies have provided evidence of moderate genetic heritability of MDD, with estimates ranging between 30-40% since these studies began in the 1980s (Coleman et al., 2020; Sullivan et al., 2000). However, single-gene candidates with high effect sizes associated with MDD in monogenic disease patterns were not found. Instead, to detect the multitude of genetic variants (single nucleotide polymorphisms, SNPs) contributing risk to MDD, genome-wide association study (GWAS) designs investigate thousands of people in a case-control or quantitative trait paradigm to detect SNPs with small-to-moderate effect sizes cumulatively contributing to probability of having the trait. The largest MDD GWAS meta-analysis to date (Howard et al., 2019) pooled GWAS data from the 2018 Psychiatric Genetics Consortium (PGC) MDD mega-analysis (Wray et al., 2018), 23andMe and the UK Biobank, with a total sample size of 807,533 individuals (246,363 MDD cases and 561,190 controls). This analysis yielded 102 variants significantly associated with MDD in 101 independent loci. Of note, significant positive genetic correlations were found between MDD and cardiometabolic traits like coronary artery disease, triglycerides, and waist-to-hip ratio, implying some shared genetic architecture of SNPs associated with MDD with these traits. The authors also found significant enrichment of SNP heritability of MDD in tissue-specific gene expression analyses within the anterior cingulate cortex (ACC) and frontal cortex, both of which are important regions for higher-order cognitive functions. The ACC has been implicated in autonomic system regulation of blood pressure and heart rate, and has a “top-down” emotional-regulation projection to both the amygdala and prefrontal cortex in response to emotional stimuli (Stevens et al., 2011). A dysregulated fronto-limbic network has been associated with MDD, with hyperactivation of regions like the ACC leading to increased attention towards and processing of emotional stimuli, with tendencies to focus more on negative stimuli (Sliz & Hayley, 2012). What is lacking from many of these large-scale analyses, however, are analyses of sex and gender differences in the shared associations between cardiometabolic dysregulation and MDD.

There are consistent gender differences in the reported prevalence of MDD in Canada and around the world, with MDD being twice as prevalent in women as it is men (Bromet et al., 2011). A study by Kendler et al. (2006) in the Swedish Twin Registry reported sex differences in heritability, with estimates of 42% in females and 29% in males, underscoring the signal of biological (sex) genetic differences (not to be conflated with gender, a social construct (CIHR, 2021)). However, few studies have been designed to be able to elucidate mechanisms of the sex and/or gender differences. We reviewed all MDD GWASs to date published in the GWAS catalog and found that only 12 (17%) of the 72 studies published conducted sex-stratified analyses (Lewis et al., 2010; Aragam et al., 2011; Shyn et al., 2011; Shi et al., 2011; Power et al., 2012; Wray et al., 2012; Ripke et al., 2013; Hyde et al., 2016; Hall et al., 2018; Zhou et al., 2018; Dunn et al., 2018; Blokland et al., 2022). In 2022, Blokland et al. conducted a sex-stratified GWAS meta-analysis of MDD in the PGC with another European regional cohort (iPSYCH) and described a SNP-heritability estimate of MDD in females almost twice (14.6%) that of males (6.6%). These authors describe a range of sex-specific genetic effects and pathways associated with MDD, primarily implicating neuronal inhibitory and excitatory regulation of brain development, but also immune and vascular pathways. The authors highlighted that the only significantly enriched pathways in the MDD gene-by-sex (G x S) gene set enrichment were in the vascular endothelial growth factor signalling pathways, a molecule which signals angiogenesis across tissue types and is also involved with vasodilation (Pandey et al., 2018).

There are many theories of how sex differences appear (Bangasser & Cuarenta, 2021), with two main overarching paradigms describing how sex differences may mechanistically arise across complex traits like MDD. There is the effect of sex hormones secreted by the gonads (ie. estrogen, progesterone, testosterone) that exert their effects on gene expression either indirectly as signal transduction molecules, or directly on the genome as transcription factors (Davey & Grossmann, 2016; Fuentes & Silveyra, 2019). Second, there are a differing number of copies of the sex chromosomes, wherein females typically have a pair of X-chromosomes, and males have an X and Y chromosome. While most genes on one of the two X chromosomes are inactivated in XX cells, higher expression of genes can occur at genes that escape X chromosome inactivation. In addition to cis-acting regulation, emerging evidence is showing how 3-D organization of the chromosomes influences gene expression and co-regulation through trans-acting mechanisms (Fleck et al., 2022), implicating autosomal regulation of genes on the X, and vice versa. However, despite the growing evidence of the importance of genes on the sex chromosomes for many diseases, the X chromosome is routinely excluded from GWAS and other genetic analyses.

A promising clinical application of GWAS is to create polygenic risk scores (PRS) to aid in primary prevention of common complex traits. For example, in population screening for common complex diseases, individuals may be identified as “high genetic risk” from their individualized PRS and receive further clinical screening and follow-up accordingly (Lewis & Vassos, 2020). Most clinical applications of PRSs have been focused on common diseases such as coronary artery disease (Klarin & Natarajan, 2022) and breast cancer (Yanes et al., 2020), and clinical trials in such areas are already underway to assess the utility of PRSs in primary prevention in addition to standard clinical risk factors. There is still much scrutiny regarding the relative economic and clinical utility of incorporating PRSs into clinical care, thus the importance of conducting analyses of group (i.e. sex) similarities and differences to inform research and clinical decisions must be emphasized and explored.

The purpose of this study is twofold: First, to investigate sex differences in the associations between MDD PRSs using sex-stratified GWAS data with MDD (and including the oft-excluded X-chromosome), and second, to compare sex differences in the relative effect sizes of MDD PRSs compared with other socioeconomic and clinical risk factors in a multivariable model of MDD. This study is unique through its use of a sex-stratified PRS methodology applied to Canadian population sample and combining genetic risk scores and cardiometabolic risk factors not commonly studied in epidemiological studies of MDD. We hypothesized that there would be genetic sex differences in the relative associations of sex-specific PRSs with MDD, and that there would be sex differences in the relative associations of the various shared sociodemographic, lifestyle and cardiometabolic risk factors associated with MDD.

## Methods

### Target Cohort Data Variable Selection: The Canadian Longitudinal Study on Aging

The Canadian Longitudinal Study on Aging (CLSA) is a longitudinal cohort study whose primary goals are to investigate the health and needs of the aging Canadian population (Raina et al., 2009). The study was designed to help understand the intersectionality of factors involved in adult development and aging, by investigating contributions of lifestyle and behaviour; psychological, biological, and clinical, healthcare services; health outcomes; and social measures (Raina et al., 2019). Since 2010, just over 51,000 Canadians aged 45 to 85 at baseline have been recruited to be followed until 2030 (or death). The CLSA is divided into the Tracking (n=21,241) and Comprehensive (n=30,097) cohorts, which vary in the type and method of data collected. Notably, only the Comprehensive cohort collected genetic data of interest to this study for 26,622 individuals.

The primary outcome variable of interest was MDD, which was encoded as a binary variable answering the question: “Has a doctor ever told you that you suffer from clinical depression?”). Sociodemographic and lifestyle covariates were chosen *a priori* because of their frequent inclusion as covariates in other epidemiological studies that investigate depression as the primary outcome variable, such as other CLSA studies (Liu et al., 2019; Shea et al., 2020; Stinchcombe et al., 2021). Self-reported sex was collected as a binary variable (male or female), total annual household income was self-reported by intervals by thousand (k) Canadian dollars in increments of <20k, 20-50k, 50-100k, >100k, and (tobacco) smoking status was collected by self-reports of never-smoker, former-smoker, or current-smoker. Three variables (alcohol consumption, highest education achieved, and partnered status) between the sociodemographic and lifestyle categories were recoded into fewer options, to provide sufficient power in downstream logistic regression analyses. Alcohol consumption frequency was recoded from 8 options to 2: “never” remained “never”; and all other options to “ever”. Highest education achieved was recoded from 6 options to 3 in accordance with Stinchome et al. (2021): The options of Bachelor’s degree or university degree or certificate above bachelor’s degree were combined as “Bachelor’s or higher”; university certificate below bachelor’s level, non-university certificate or diploma from a community college, and trade certificate from a vocational school or apprenticeship were combined to “Under a bachelor’s”; and no-post-secondary degree, certificate or diploma remained the third option “No post-secondary education”. Lastly, in accordance with Liu et al., (2019), partnered status was recoded into a binary response from five options of single, widowed, divorced, or separated to “non partnered” and married or living with a partner in a common-law relationship to “partnered”.

Clinical variables commonly studied in metabolic syndromes and/or those that exhibit notable sex differences were included as covariates. Waist-to-hip ratio (WHR) was a physical measurement ascertained in the study home visit. WHR was included in lieu of BMI, as BMI is an imprecise measure that does not account for muscle mass and/or distribution of mass, specifically regarding higher risk of metabolic disease in association with higher visceral fat (abdominal core area) compared to other areas of lower relative risk (ie. breasts and hips) (Lotta et al., 2018). Second, since there was no dyslipidemia field captured, this study aggregated data from the extensively detailed baseline medication data collected by the CLSA investigators during home visits to produce a composite yes/no binary variable “on lipid-lowering therapy?” if they were on any of the following cholesterol-lowering medications: Atorvastatin, rosuvastatin, simvastatin, lovastatin, cerivastatin, fluvastatin, pitavastatin, pravastatin alirocumab, evolocumab, ezetimibe (Dennis et al., 2021). Additionally, the binary composite variable “ischemic heart disease outcome” (IHD) was assigned to participants who self-reported having had a myocardial infarction (MI) and/or angina. The rest of the self-reported binary (yes or no) variables included were high blood pressure, diabetes, and hypothyroidism.

Chi-squared tests of independence were conducted to assess if there were significant differences by MDD status in the frequencies of the options for each categorical variable. For the numeric variables, means and standard deviations were calculated and statistical significance between MDD and no-MDD (within-sex) means were assessed via two-sample t-tests.

Correlation matrices with associated p-values were generated using the corrplot package (Wei, 2021), with covariates mapped against each other. Additionally, covariates were retained that did not have high correlation (Pearson r >> 0.5) with other covariates.

### CLSA: Quality Control and Filtering of Genetic Samples

Genome-wide genotyping data were collected from DNA extracted from the buffy coat of blood samples and genotyped using the 820K UK Biobank Axiom Array (Affymetrix), consisting of 795,409 genotyped SNPs for each of 26,622 genotyped individuals, and were received in Plink 1.9 format (Chang et al., 2015). Quality control (QC) tests were conducted following thresholds described in an accompanying Genome-wide Genetic Data Release documentation (Forgetta et al., 2022).

Samples were excluded based on four criteria: i) Having at least one relative of 3^rd^ degree or closer among the set of genotyped individuals (n = 1637), wherein relatedness was determined using KING software program version 2.1.3 (Manichaikul et al., 2010), which assesses relatedness by first computing kinship coefficients, then analyzing the proportion of identity-by-descent (IBD) to the coefficient. ii) Extreme outliers in sample-wise heterozygosity were excluded, wherein extreme values suggest low quality genotyping or cross-contamination of biological samples (n = 15 removed). Outliers were determined using Plink’s “het” flag that generated a threshold value related to the proportion of the homozygous and heterozygous frequencies. iii) Sex-mismatched samples (n = 33) and samples with missing sex chromosomal data (n = 15) were removed. These values were determined by when the self-reported gender did not match the Plink-determined chromosomal sex, or if Plink could not assess sex and the sample was thus labeled as “ambiguous”. iv) Participants were filtered by genetic ancestry to retain European-ancestry individuals only. These individuals had been assigned “PCA cluster ID 4” in the sample QC text file by Forgetta et al. (2022) and filtered accordingly (n = 1963 non-European samples removed). To obtain these cluster ID values, the CLSA data analysts conducted population structure analyses via principal component analyses (PCA) by comparing the CLSA participants to 414 individuals across 4 superpopulations (Western European, Han Chinese, Japanese and Yoruban) from the 1000 Genomes phase 3 reference population using k-means clustering. The top 10 principal components (PCs) for the CLSA European ancestry subset were provided and integrated in downstream analyses.

The SNP QC was conducted on the genotyped markers on the European-ancestry subset, as described by Forgetta et al. (2022). The retained markers met four main criteria within this subset and were flagged accordingly in the accompanying SNP QC text file. Markers were flagged by CLSA analysts under the following conditions: i) Extreme discordance from expected genotype frequency between batches (p < 3.15*10^−10^) (n = 14,753); ii) Deviation from Hardy-Weinberg Equilibrium (p<10^−6^) for diploid regions of the genome (autosomal and pseudo-autosomal regions (PARs) of the X/Y chromosomes) (n = 7,790); iii) <0.99 Discordance between the sample and 4 control replicates on each genotyping plate(n = 27,937). iv) Genotype frequency discordance stratified between chromosomal sexes (female 46, XX and male 46, XY) below Fisher’s p < 3.15*10^−10^ (n = 248). This statistic was calculated using Fisher’s exact test on the 2×3 table of genotype counts, or 2×2 table of allele counts for the sex-specific, non-PAR region of the X chromosome. v) insertions and deletions (“indels”) (n = 15,616), so as only single-character allele codes of A, C, G or T were retained. vi) low frequency SNPs of MAF < 0.01 (n = 95,363). Slight differences in the final number of SNPs between males, females and both-sexes in the autosomes and X-chromosomes are negligible, and most likely due to sampling artefact and not reflective of true sex differences.

### Base MDD GWAS: UK Biobank Summary Statistics

The UK Biobank (UKB) is a prospective health study of over 500,000 individuals from the UK, recruited between ages 40 and 69 at recruitment (2006-2010) (Bycroft et al., 2018). Like the CLSA, this is a “representative” population cohort, such that there isn’t a particular bias or enrichment by a particular trait or disease. The most reported ethnicity was British (88.26%), which maps to ancestral superclusters of European genetic ancestry when compared to the 1000 Genomes phase III (Bycroft et al., 2018). MDD was encoded from self-reported history of past or present (clinical) depression through the question “has a doctor ever told you that you have depression” (phenotype code 20002_1286). The genotyping array created by the UKB, the UK Biobank Axiom® (ThermoFisher Scientific, 2014), was the same array used by the CLSA cohort in genotyping their participants.

For UKB self-reported history of clinical MDD, three sets of summary statistics from GWASs of sex-stratified and non-stratified analyses were downloaded from the Neale Lab Website (Neale Lab^a^, 2018). The non-stratified (“both-sexes”) GWAS had a combined sample size of 361,141, the female GWAS had a sample size of 194,121, and males had 167,020 (all included data for 13,791,467 SNPs). The Neale Lab reported 20,648 cases and 340,493 controls retained in their overall (both-sexes) analyses, with a trait prevalence for self-reported depression of 5.72% (Neale Lab^b^, 2019). The number of cases and controls for their sex-stratified analyses are not publicly provided on the Neale Lab website.

Each of the both-sexes, female-only and male-only UKB MDD GWAS summary statistics data were processed through a publicly-accessible quality control pipeline developed in-house (Belikau, 2021). This QC excluded SNPs that did not have a p-value within (0,1), didn’t have a valid SNP rs-id, the chromosome value didn’t match human, the base pair wasn’t an integer, one of the alleles was not a single nucleotide variant (ie. an insertion), if the SNP was palindromic, and if the SNP wasn’t biallelic. Duplicated rows were identified and removed in R v4.0.2 (R Core Team, 2017). In sum, 11,937,067 SNPs remained post-QC (86.6% of input) for the both-sexes GWAS, 11,935,604 SNPs (86.5%) for the female-only GWAS, and 11,934,922 SNPs (86.5%) for the male-only GWAS.

SNP heritability (*h^2^*_SNP_) estimates and genetic correlation analyses were conducted using linkage disequilibrium score regression software LDSC (Bulik-Sullivan et al., 2015). The outputs of this software are the estimates of MDD *h^2^*_SNP_ of the input GWAS summary statistics, as well as the genetic correlation (r_g_) between the female- and male-only GWASs.

### Polygenic Risk Scores

Polygenic risk scores for the CLSA data were constructed using the software PRSice-2 (Choi & O’Reilly, 2019), in which a clumping and thresholding (C+T) method was applied on SNP summary statistics from base UKB GWAS data. Clumping (“C”) refers to the iterative selection of the most significant SNP of a group in linkage disequilibrium. Thresholding refers to the removal of SNPs with trait-associated p-values above a certain significance threshold (“T”). For the remaining SNPs after C+T, PRSice-2 calculates a PRS for each individual (*j*) with the equation below, in which *S* is the effect estimate for the effect allele (*i*), and *G* is the dosage of the effect allele (0, 1 or 2).

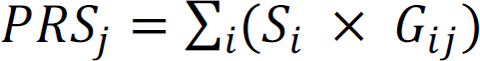

PRSice-2 does not currently permit sex-chromosome input as chromosome 23. As such, X-chromosome SNP-genotyped data were extracted separately from the autosomal data and provided a new chromosomal label “22” so as to be recognized by PRSice-2. This did not compromise or change the encoding within the original CLSA .bed file wherein the X-SNPs were already encoded as 0/1/2 for females and 0/2 for males. PRSice-2 was run on these data under additive model assumptions for both sexes, resulting in X-PRSs to be assessed separately from the autosomal-PRSs. Autosomal PRSs for each sex were created from the sex-specific (male or female) GWAS, and a PRS from the sex-specific X-chromosome data. Each PRS had the same C+T parameters set to p-value threshold of p<0.01, LD-clumping distance of 250 kilobases (kb), and clump-r^2^ of 0.1.

CLSA participants were sex-stratified into female- and male-only groups, to which PRSs created from base GWAS data that were either sex-stratified (hereafter “female-only” and “male-only”) or data from both sexes were pooled (hereafter “both-sexes”). If there are indeed differences, ie. of a PRS made from male-only MDD GWAS data tested on females shows no associations with female MDD, and vice versa, then this implies there may be sex-specific genetic associations missed when sex is regressed out as a covariate in the GWAS analyses. The purposes of these analyses were thus to test this if the sex-specific and both-sexes PRSs were transferable between the subsetted samples.

Individuals were labeled by decile in which their score fit relative to the rest of their sex-specific group. A univariable logistic model was performed between the PRS deciles and MDD, with the first (0-10) decile label being the referent value, and estimates converted to odds ratios using the broom package (Robinson et al., 2022) in R v4.0.2.

### Multivariable Model

A multivariable logistic model was fit for genetic, sociodemographic, lifestyle and clinical variable associations with an MDD outcome, using the *glm* function in R 4.0.2. Using the *broom* package (Robinson et al., 2022), multivariable model output estimates were exponentiated into odds ratios and 95% confidence intervals. There were two main goals of this modelling: first, to investigate which variable(s) had significant effect sizes (converted to odds) with MDD in the multivariable model, and second, to investigate the relative proportion of how much each variable contributed to the model. As such, the model was as follows:

> MDD ∼ Sex-specific Autosomal PRS + Sex-specific X-PRS + Age + Partnered Status + Annual Income + Highest Education + Smoking Status + Frequency of Alcohol Consumption + LLT + WHR + High blood pressure + Diabetes + Hypothyroidism + IHD

Prior to input, all numeric variables (PRSs, age, waist-to-hip ratio) were scaled to a mean of zero and standard deviation of 1. Logistic regression models were then conducted to investigate the relative proportion of each variable and variable category’s pseudo-R^2^ contribution to the association with MDD by comparing pseudo-R^2^ values using the *rms* package (Harrell, 2021) in R v.4.0.2. There were two sets of analyses: the first goal was to compare the relative contribution of each covariate category to the total model of associations with MDD, the second being the individual covariates alone.

## Results

### Base MDD GWAS: UK Biobank Results

Manhattan plots for the three UKB summary statistics data post-QC were made in R v4.0.2 using the *gwaRs* package (Nkambule, 2021), with SNPs that passed genome-wide significance of p < 1*10^−8^ depicted in red, shown in Figure 1. Corresponding quantile-quantile (Q-Q) plots were made with the *fastman* R package (Hwang, 2019), representing the distribution pattern of SNP p-values. As the genomic inflation factors (λ) for all plots were under 1.1, there is not likely to be structural bias (e.g., population structure) within each of the GWAS to cause inflation. However, as there are a lot of deviations of the expected versus observed p-values as shown in the Q-Q plots, this is strongly indicative of trait polygenicity. The UKB SNP-heritability results were female h^2^_SNP_ = 0.02 (se = 0.0026), male h^2^ = 0.013 (se = 0.0028), and their genetic correlation was r_g_ = 1.072 (se = 0.167, p = 1.34e-10).

**Figure 1:**
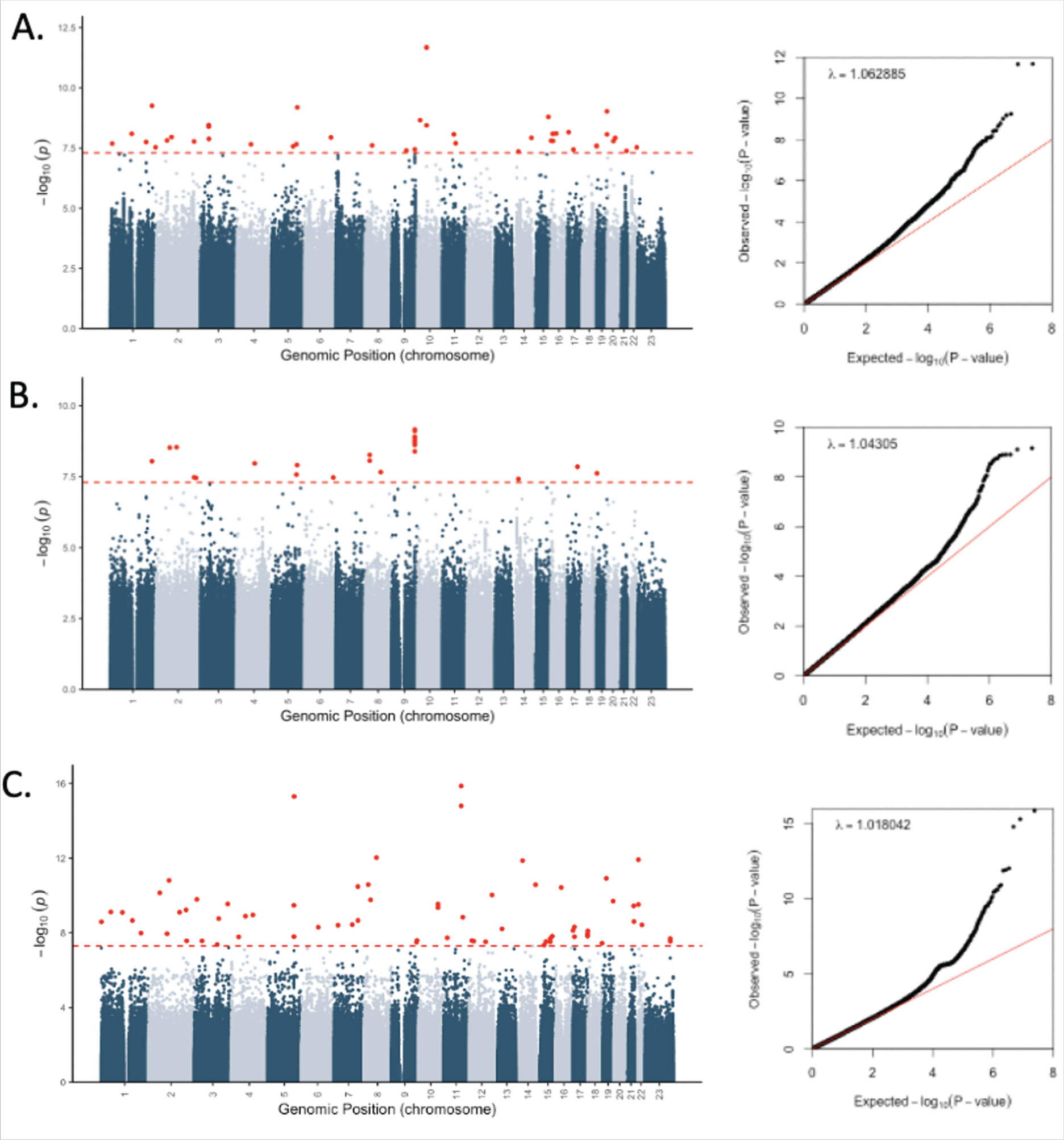
Manhattan plots (post-QC) of the UK Biobank self-reported depression (phenotype code 20002_1286) with their associated QQ plots. Red line in Manhattan plots represents genome-wide significance (p < 1*10-8), and the red line in the QQ plot demonstrates y=x. λ represents the calculated genomic inflation factor. A) Both-sexes data (not stratified), n=361,141; B) Female-only, n=194,121; C) Male-only, n=167,020.

### Sex Differences in PRS Associations with MDD

Results in Figure 2 demonstrate odds of having MDD per PRS decile with 95% confidence intervals for the autosomes for all three groups (females, males, both-sexes). PRSs were made for the X-chromosome with dosage differences accounted for with males encoded as 0/2 and females 0/1/2, however results in decile comparisons were not significant. Exact values of these autosomal and X-chromosome associations are reported in Supplementary Table 1. Within the female-only data (Figure 2A), the PRS made from female-specific GWAS outperformed the PRS made from the both-sexes data, with higher odds across all deciles. Within the male-only data (Figure 2B), however, the male sex-specific PRS did not perform better than the both-sexes data.

**Figure 2:**
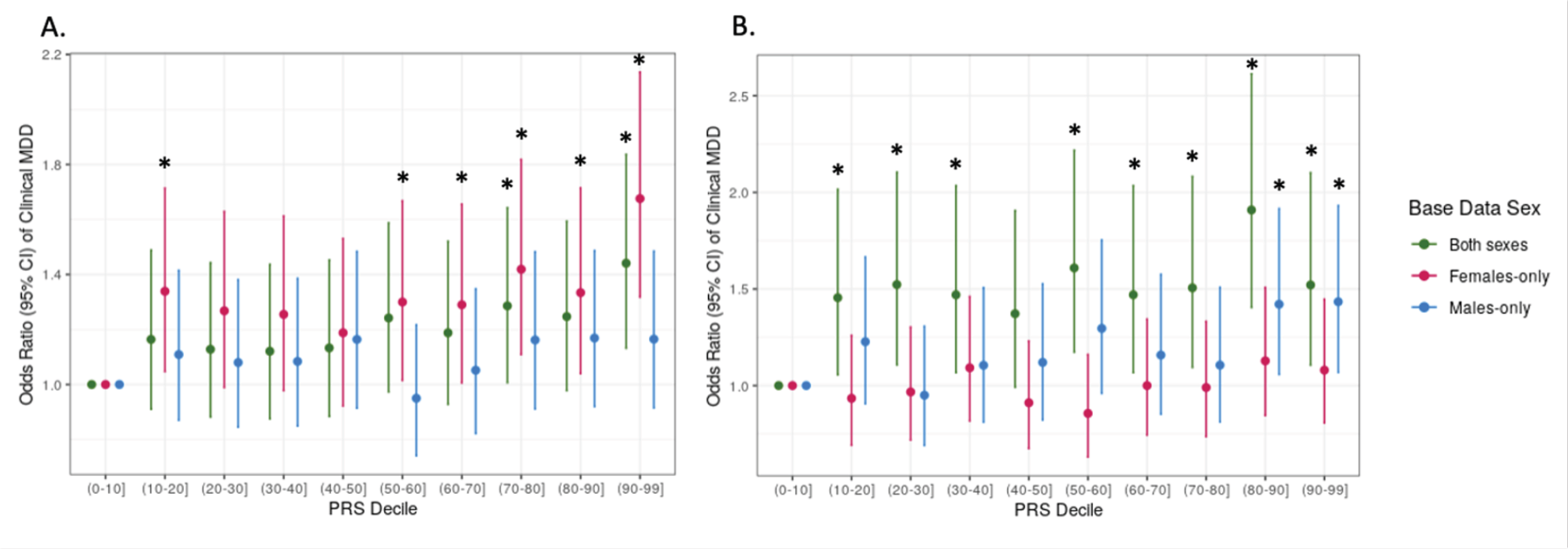
Odds ratios with 95% CI of MDD with each increase in PRS decile across target data. Asterisks (*) denote significant difference in odds of MDD by PRS decile compared to the the referent (lowest) decile (p<0.05). Colours represent different UKB base GWAS data used to make each PRS: green was the both-sexes GWAS, magenta the female-only GWAS, and blue the male-only GWAS. A) PRSs applied on target autosomal data of females in the CLSA. B) PRSs applied on target autosomal data of males in the CLSA.

### CLSA: Exploratory Data Analyses

Following genetic analyses, other non-genetic variables were investigated under a sex-stratified framework to elucidate independent and relative associations with MDD to each other. Filtering of samples with NAs in any of these categories, summarized in supplementary Table 2, removed 6,097 samples further from the 22,974 from sample QC. The final model post-filtering resulted in 8,207 females (6,471 without MDD (79%) and 1,741 with MDD (21%)) and 8,670 males (7,615 without MDD (88%) and 1,055 with MDD (12%)) for a total of 16,877 participants remaining. Table 1 outlines sex-stratified covariate summary statistics for individuals with a history of self-reported clinical MDD compared to those without MDD.

**Table 1.** Final counts and summary statistics of all categorical (frequency/%) and numerical (mean/SD) variables. These counts reflect participants who were retained after PRS QC steps and covariate QC that are reflected in multivariable analysis (figure 4.8). For categorical variables, chi-squared tests of independence for p-values between MDD vs. non-MDD groups were calculated (within-sex comparisons), and for numerical variables, p-values were calculated using 2-sample t-tests.

|  |  |  | Females (n=8,207) |  |  | Males (n=8,670) |  |  |
| --- | --- | --- | --- | --- | --- | --- | --- | --- |
|  | Variable | Label | Without MDD<br>(n=6,471) | With MDD<br>(n=1,736) | P | Without MDD<br>(n=7,615) | With MDD<br>(n=1,055) | P |
| **Sociodemographic** | Age (mean, SD) |  | 61.9 (10) | 59.9 (8.7) | 3.00E-15 | 62.7 (10.1) | 60.7 (8.7) | 2.52E-09 |
|  | Partnered status (count, %) | Partnered | 4202 (65) | 923 (53) | < 2.2E-16 | 6167 (81) | 766 (73) | 1.87E-10 |
|  |  | Not partnered | 2269 (35) | 813 (47) |  | 1448 (19) | 289 (27) |  |
| | Annual income (count, %) | >\$100k | 2491 (38) | 518 (30) | < 2.2E-16 | 3720 (49) | 422 (40) | < 2.2E-16 |
| | | \$50-100k | 2377 (37) | 601 (35) | | 2715 (36) | 369 (35) | |
| | | \$20-50k | 1388 (21) | 470 (27) | | 1026 (13) | 202 (19) | |
| | | <\$20k | 215 (3) | 147 (8) | | 154 (2) | 62 (6) | |
|  | Highest education (count, %) | Bachelors or higher | 3236 (50) | 852 (49) | 6.05E-01 | 4432 (58) | 607 (58) | 8.81E-01 |
|  |  | Some post-secondary education under a bachelor's | 2651 (41) | 715 (41) |  | 2555 (34) | 357 (34) |  |
|  |  | No post-secondary education | 584 (9) | 169 (10) |  | 628 (8) | 91 (9) |  |
| **Lifestyle** | Smoking status (count, %) | Never smoker | 3429 (53) | 803 (46) | 3.98E-10 | 3583 (47) | 436 (41) | 1.80E-10 |
|  |  | Former smoker | 2632 (41) | 756 (44) |  | 3427 (45) | 472 (45) |  |
|  |  | Current smoker | 410 (6) | 177 (10) |  | 605 (8) | 147 (14) |  |
|  | Frequency of alcohol consumption (count, %) | Never | 611 (9) | 235 (14) | 6.29E-07 | 595 (8) | 157 (15) | 2.10E-14 |
|  |  | Ever | 5860 (91) | 1501 (86) |  | 7020 (92) | 898 (85) |  |
| **Clinical** | Lipid-lowering therapy (LLT) (count, %) | Not on LLT | 5694 (88) | 1471 (85) | 2.95E-04 | 5809 (76) | 790 (75) | 3.17E-01 |
|  |  | On LLT | 777 (12) | 265 (15) |  | 1806 (24) | 265 (25) |  |
|  | Waist-to-hip ratio (mean, SD) |  | 0.83 (0.07) | 0.84 (0.07) | 1.40E-08 | 0.97 (0.06) | 0.98 (0.07) | 3.37E-10 |
|  | High blood pressure (HBP) (count, %) | No HBP | 4450 (69) | 1159 (67) | 1.11E-01 | 4810 (63) | 593 (56) | 1.24E-05 |
|  |  | HBP | 2021 (31) | 577 (33) |  | 2805 (37) | 462 (44) |  |
|  | Diabetes (count, %) | No diabetes | 5639 (87) | 1412 (81) | 6.61E-10 | 6336 (83) | 818 (78)% | 5.55E-06 |
|  |  | Diabetes | 832 (13) | 324 (19) |  | 1279 (17) | 237 (22) |  |
|  | Hypothyroidism (count, %) | No hypothyroidism | 5291 (82) | 1323 (76) | 2.03E-07 | 7139 (94) | 974 (92) | 7.65E-02 |
|  |  | Hypothyroidism | 1180 (18) | 413 (24) |  | 476 (6) | 81 (8) |  |
|  | Ischemic Heart Disease (IHD) Outcome (count, %) | No IHD | 6264 (97) | 1651 (95) | 6.98E-04 | 6958 (91) | 938 (89) | 8.57E-03 |
|  |  | IHD | 207 (3) | 85 (5) |  | 657 (9) | 117 (11) |  |

Correlations between covariates were assessed to determine exclusion of variables and identify relationships between covariates that exhibited sex-specific trends compared to pooled both-sexes data. Correlation heatmaps are shown in Figure 3. Despite several variables indicated with asterisks (*) that had significant correlations, Pearson’s r coefficients were < 0.5 and were thus justified in their retention in the model per *a priori* hypotheses. Corresponding correlation values (-1 to 1) and p-values matrices are in Supplementary Tables 3-8.

**Figure 3:**
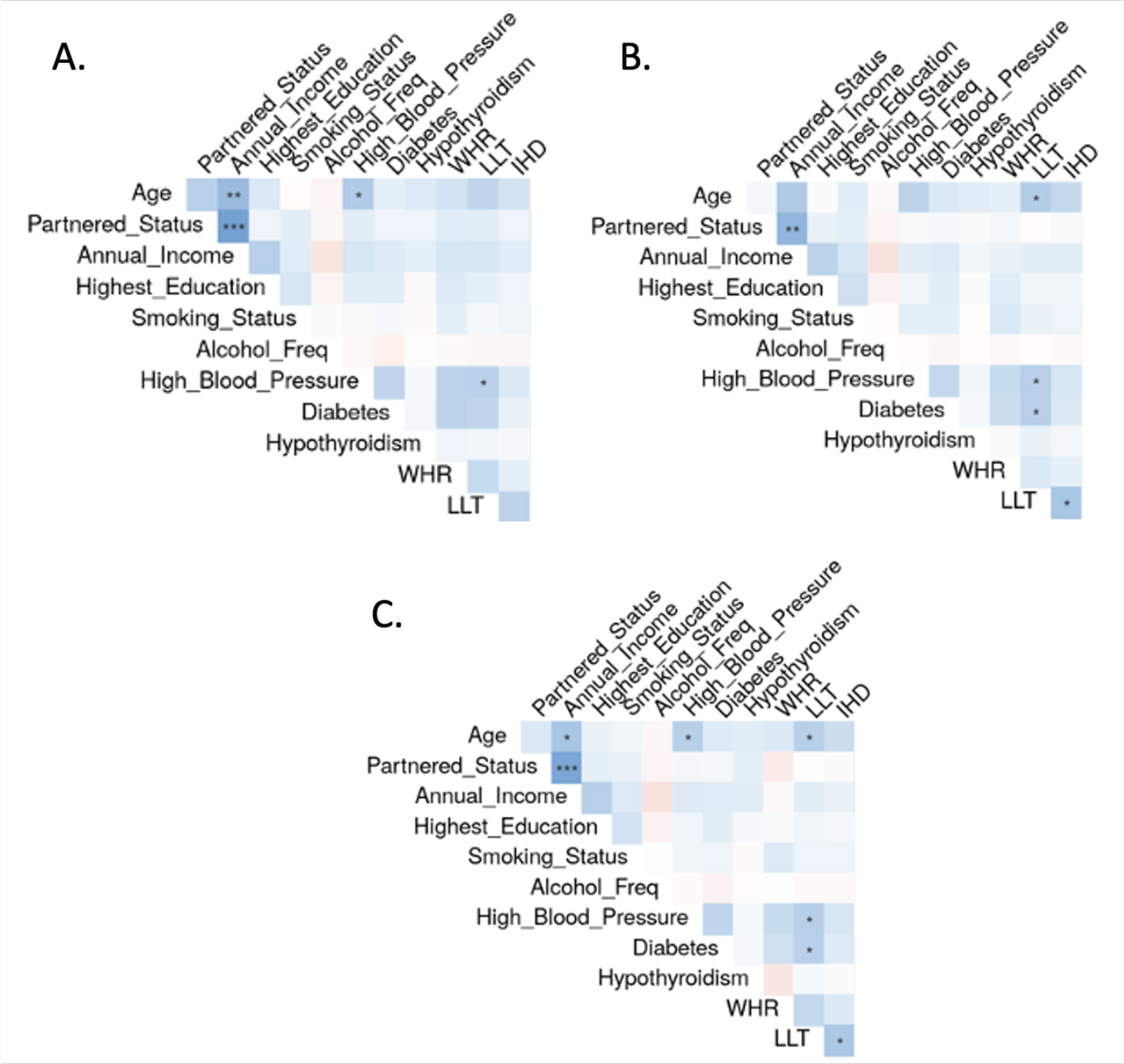
Correlation heatmap of covariates within the CLSA. Asterisks denote significance of p<0.001 (***), p<0.01(**) and p<0.05(*) between variables within sex-stratified or non-stratified data. A: Female correlation matrix; B: Male correlation matrix; C: Both-sexes correlation matrix.

Overall, most covariates were not significantly correlated with each other. Few covariates were significant in both sexes in the sex-stratified matrices (Figure 3A and B), such as high blood pressure with lipid-lowering therapy (LLT) (males’ r = 0.25, p = 0.037 and females’ r = 0.24, p = 0.043))). There were some observed sex differences in some variables that were significantly correlated in one sex but not the other (ie. age and high blood pressure; males’ r = 0.23, p = 0.071 vs. females’ r = 0.27, p = 0.032). More variables in the male correlation matrix significantly correlated with LLT (age, HBP, diabetes, IHD) than in females (HBP only).

### Multivariable Modelling

Multivariable logistic modelling results are visualized in forest plots in Figure 4. Males and females overall were very similar and often showed significant associations with similar direction of increased or decreased odds of MDD in sociodemographic variables.

**Figure 4:**
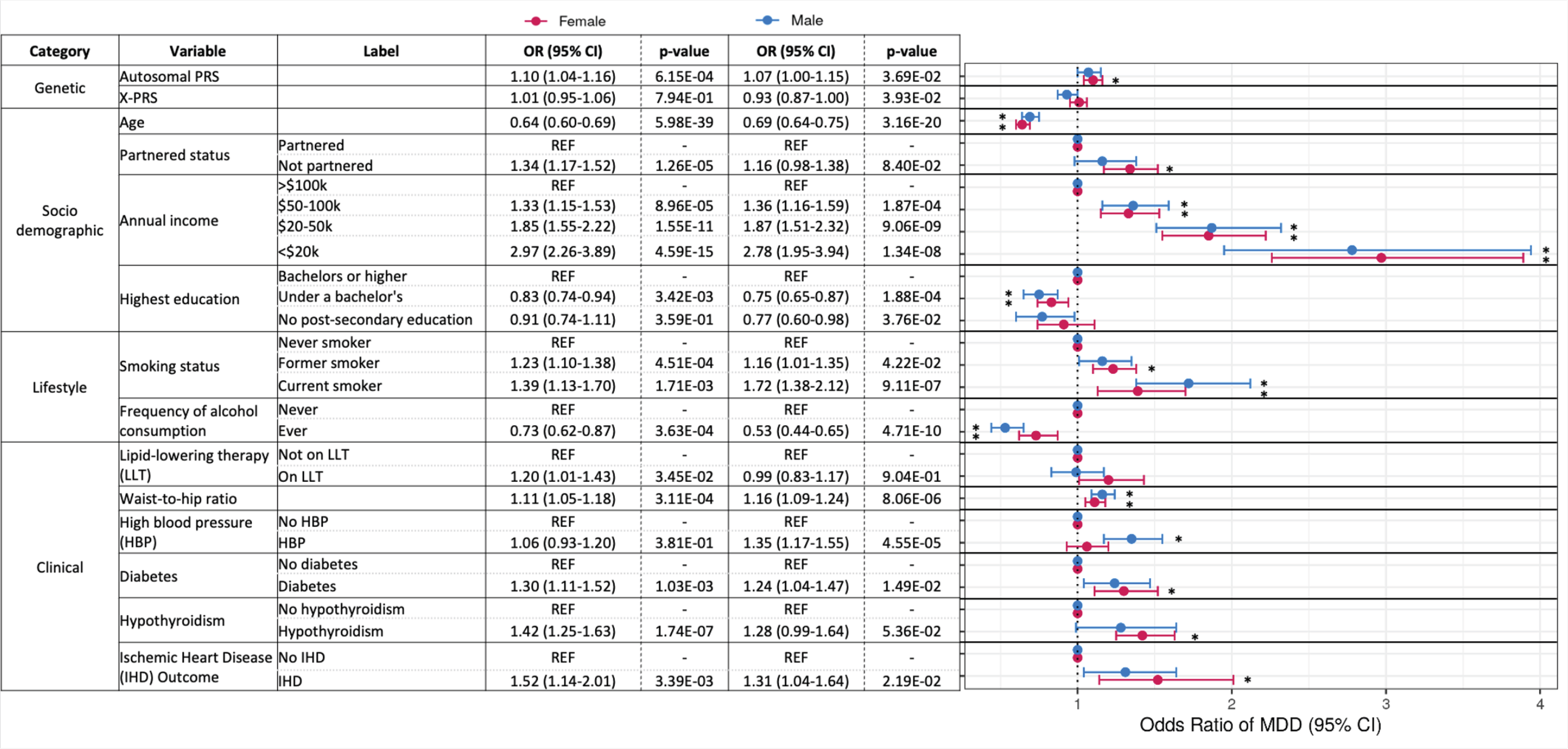
Odds of MDD (95% CI) for sociodemographic, lifestyle and clinical covariates, stratified by sex. Sex-specific autosomal base data were used for generating the autosomal and X-PRSs. Asterisks (*) represents significance after adjusting for multiple testing (p<0.05/14) compared to the referent (REF) value.

Age had the highest effect size and significance in associations with MDD among all variables included in the multivariable models, with younger age associated with higher odds of MDD for both sexes (females OR = 0.64 (0.60-0.69), males OR = 0.69 (0.64-0.75)). Next most significantly of all variables, lower annual income associated with higher relative odds of MDD for both sexes (female OR = 2.97 (2.26-3.89) and male OR = 2.78 (1.95-3.94)). A sex-specific association with partnered status was observed, with being unpartnered associated with higher odds of MDD, but only in females (OR = 1.34 (1.17-1.52)). Current or former smoking status was associated with higher odds of MDD for females, but just current smoking status for males. However, there was a higher relative effect size for current males (OR = 1.78 (1.42-2.21)) than females (OR = 1.38 (1.12-1.69)), although not significant, male and females if they were former smokers (OR = 1.24 (1.10-1.39). For both sexes, people who reported never drinking alcohol had higher odds of MDD than people who ever drank alcohol, with a larger effect observed in males (OR = 0.53 (0.44-0.65)) than females (OR = 0.73 (0.62-0.87)).

For the clinical variables, there were sex differences in almost every variable that was significant. The only variable that was significant for both sexes was waist-to-hip ratio (WHR), in which higher odds of MDD was associated with an increase in each scaled unit (female OR = 1.11 (1.05-1.18), male OR = 1.16 (1.09-1.24)). Having high blood pressure was associated with higher odds of MDD in males (OR = 1.38 (1.19-1.60)), but not females. For females only, they had higher odds of having MDD if they have diabetes (OR = 1.30 (1.11-1.52)), hypothyroidism (OR = 1.42 (1.25-1.63) and/or a history of IHD (OR = 1.52 (1.14-2.01).

Being on lipid-lowering therapy did not pass significance in the multivariable model of MDD for either sex. When comparing LLT and IHD frequency in this “healthy” cohort, between 2-3 times more participants were on LLT (female n = 1,984, male n = 3,382) than participants with a IHD outcome (female n = 594, male n = 1379). However, of the 1,984 females and 3,382 males taking LLT, the majority did not have a history of IHD (female n = 1,674 (84%), male n = 2,469 (73%). This may interpreted such that these participants could be on preventive medical management against cardiovascular disease outcomes, which is unsurprising given the age range of this cohort (45-85 years).

These results of covariates contributing significantly to the odds to MDD paint a complex picture of MDD that supports the hypothesis that there is a cardiometabolic component to MDD, and that there are some sex differences in which systems and dysfunctions associate more (or less) with MDD.

### Variable Contributions to MDD: Pseudo-R^2^

The relative proportions of the pseudo-R^2^ for all covariate categories (Figure 5A) and individual covariates that contribute to the female and male MDD models (Figure 5B) show sex differences in relative contributions of different variables and variable categories to MDD. Pearson’s chi-squared test (conducted in R v4.0.2) confirms significant differences between the sexes when examining these trends across variable categories (p = 0.002) or individual variables (p = 0.005). These data are summarized in Supplementary Table 9 for both the variables and variable categories. The total pseudo-R^2^ for the female model for MDD was 0.070, which was slightly higher than the male model (pseudo-R^2^ = 0.058).

**Figure 5:**
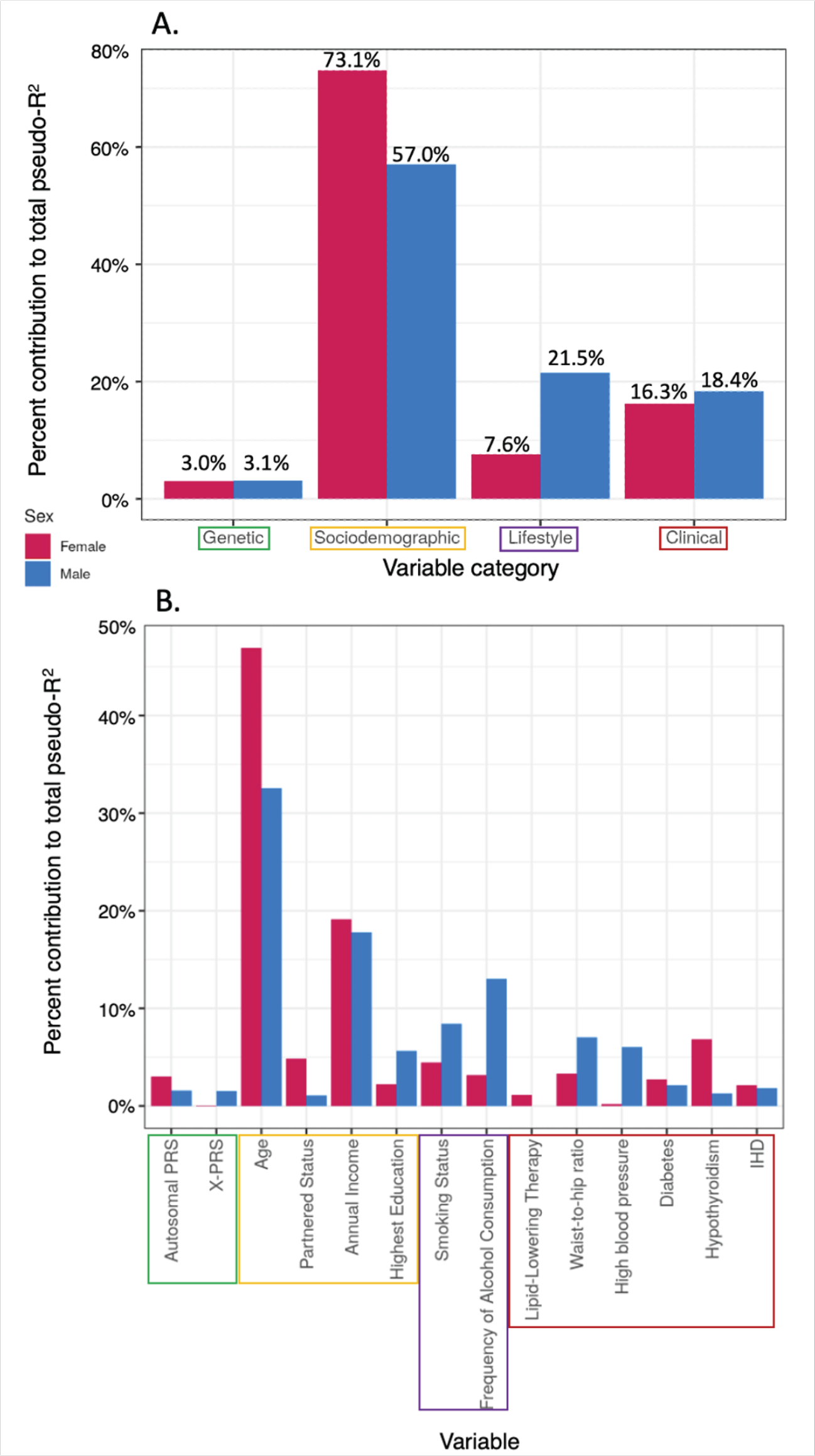
Sex differences in relative proportions of variables’ contributions to MDD logistic model’s pseudo-R^2^. Autosomal and X-PRS were both made with sex-specific GWAS base data. Bars represent a relative proportion (%) of Pseudo-R^2^ contributed to the model. Pearson’s Chi-squared tests were conducted to evaluate significant differences between the sexes across all: A) variable categories (p=0.002), or B) individual variables (p=0.005).

As shown in Figure 5A, variables in the sociodemographic category contribute the majority to both the female model and to a lesser extent, the male model. The distribution of the rest of the variables’ contributions to model are quite different between categories, with more of the male model of MDD represented by the genetic, lifestyle and clinical categories compared to females. This begets the question: which variables within each of these categories are explaining the majority of variance in this model, and are there sex differences? Figure 5B addresses this question, with visibly different proportions of each covariate’s contributions to the MDD model for each sex. The five covariates contributing the highest proportion towards the variance of the model for females were age (46.9%), annual income (19.1%), hypothyroidism (6.8%), partnered status (4.9%), and smoking status (4.5%), totalling 82.2% of the full female model. For males, the top five were age (32.5%), frequency of alcohol consumption (13.0%), annual income (17.8%), smoking status (8.4%) and waist-to-hip ratio (7.1%), totalling 78.9% of the full male model. Lastly, there are stark sex differences in the magnitude of the autosomal and X-PRSs’ contributions, despite contributing relatively little to the overall models. In females, the female-specific autosomal PRS contributed 3.0% of the category’s total 3.02%, with the X-PRS contributing close to 0 (0.02%). In contrast, the male-specific autosomal and X-PRSs each contributed half (1.59% and 1.55%, respectively) to its category’s total (3.14%) in males.

Altogether, these analyses suggest that MDD PRSs are not covariates that contribute large effect sizes to models of MDD inclusive of other epidemiological covariates. However, these models do suggest that including cardiometabolic risk factors (ie. high blood pressure, waist-to-hip ratio, diabetes, hypothyroidism) not traditionally included in studies of MDD may be of utility in future studies of MDD with available clinical data.

## Discussion

In sum, there are both genetic and epidemiological sex differences of risk factors associated with higher odds of MDD in the CLSA. The sex-specific polygenic risk scores for MDD contributed a negligible effect size to MDD risk in the multivariable model compared to sociodemographic (ie. age) and cardiometabolic risk factors. These results underscore the need for increased attention to study design and analysis paradigms that allow for sex and gender differences to be elucidated in associations with complex traits such as MDD.

A motivation for investigations into PRSs is their potential clinical utility, as the mechanistic insights gained from GWAS studies can inform risk stratification and prevention for complex diseases. Many have expressed concern over the generalizability and accuracy of these PRSs between diverse groups of people, be it genetic ancestries, age, sex and/or genders, such that these scores have questionable utility in clinical practice (Wand et al., 2021; Wray et al., 2021). Therefore, despite the progress in psychiatric genetics in improving PRS prediction methods within and across traits, a common sentiment is that there is much to be improved before these genetic risk scores can be integrated into routine care (Murray et al., 2020). Regarding biological sex, the results of this study suggest that the female-specific MDD PRS captures more of the genetic risk of MDD in females than the male MDD PRS does in males. One interpretation of this could be phenotyping gender bias in the UKB and/or CLSA, meaning males may not have been as accurately phenotyped as females. A second interpretation could be that there is a slightly higher genetic contribution of the SNPs in the female PRS towards odds of MDD than the SNPs in the male PRS towards male MDD.

The lack of GWASs including the X-chromosome to account for X-chromosome inactivation and dosage differences by sex is a major limitation in the field to understanding a portion of sex differences due to the chromosomal complement, despite the availability of statistical methods (König et al., 2014; Wise et al., 2013) and software (Chang et al., 2014; Gao et al., 2015) to do so. X-PRSs were made with the UKB X-chromosome GWAS data, which included a genome-wide-significant peak on the X chromosome in males, but not in females. In this study, the lack of significant differences between X-PRS-derived risk deciles does not rule out the importance of including the X-chromosome in future GWASs and downstream analyses, such as gene-gene (G x G) and gene-by-environment (G x E) analyses: investigating the genes on the X-chromosome in conjunction with the autosomes (the other “genes”) and comorbid metabolic syndromes or socioeconomic situations that may be stressors (the “environment”) could still lead to vital insights in future studies. Additionally, studies that exclude the X are forgoing important genes encoded on this chromosome with well-established roles in endocrine and immunometabolic processes, such as the androgen receptor (AR), the angiotensin-converting enzyme 2 (ACE2) in blood pressure regulation, and toll-like receptors (TLR) 7 and 8 with roles in innate immune system function.

Sex-specific MDD risk was observed in association with different cardiometabolic variables, and a much larger portion of variance in the model was driven by age in females than in males. The results of the multivariable model underscoring age as the majority contributor to the model’s variance for women – in this cohort where baseline age was 45 – suggests that there may be a pre- or peri-menopausal effect driving the association with higher odds of MDD in the younger cohort. Future studies warrant inclusion of female-specific variables (ie. menopause status, hormone therapy use, etc.) in sex-specific analyses to disentangle a hormonal association with MDD and potential interaction with the PRSs.

The complex intersection of shared behavioural and metabolic adaptions to chronic stress does not imply a shared pathway, but rather a complex, interconnected network of biological liability between the traits with chronic inflammation and homeostatic dysregulation (Harshfield et al., 2020; Khawaja et al., 2009; Tobaldini et al., 2020). In particular, consequences of maladaptive responses to stress (ie. smoking, poor diet and exercise) have complex interactions with biological systems, which in turn influence risk of MDD and IHD through chronic activation and resulting consequences of the HPA and HPG axes (Tobaldini et al., 2020). There are sex differences in how the inflammatory response to stress is modulated, particularly via interactions of the magnitude and ratio of circulating sex hormones (specifically, estradiol to testosterone). For example, in the brain, activated estrogen-receptor alpha (ERα) results in inhibition of intracellular transport of signaling molecules induced by inflammatory stimuli (Vegeto et al., 2008). Estrogens have immuno-enhancive effects; in contrast, testosterones have immunosuppressive effects, via interactions within B- and T-cell maturation (Taneja, 2018). For example, estrogens have been shown to have anti-hypertensive effects through interactions with key effector molecules (angiotensinogen and renin) in the vasoconstrictor/vasodilator system of blood vessels (Medina et al., 2020). This underscores the need to consider the effect of sex hormones and relative concentrations over the life cycle in future sex-disaggregated studies.

Although decreased estrogens post-menopause may be associated with lower risk of MDD compared to pre-menopausal levels, if women experience myocardial infarction (MI), they are significantly more likely than men to experience depression and anxiety post-MI (Liblik et al., 2021). Recent evidence suggests that MDD precedes CAD risk more than CAD precedes MDD risk for both sexes, although MDD has been shown to associate more with CAD in women than men (Honigberg et al., 2022). Higher genetic risk for MDD in women has recently been shown to associate with higher risk for cardiovascular disease in women compared to men (Jiang et al., 2022). This literature supports results from this study showing females’ increased risk for MDD with comorbid diabetes, hypothyroidism and/or a previous IHD diagnosis, without the same trends observed in males.

### Limitations

MDD phenotype ascertainment is an ongoing challenge in the literature when comparing and contrasting cohort profiles. The CLSA is a “representative” Canadian population cohort who were older (over 45) at recruitment. As detailed psychiatric assessments of MDD were not integrated in the study design, there are limitations in the reliability (ie. stringency of depression, MDD phenotype) of the phenotype ascertainment. Future analyses requiring MDD phenotyping of large biobank cohorts may benefit from integrating multiple depression measures available to more accurately assess MDD status, rather than choosing a single measure, as demonstrated by Glanville et al (2021). Moreover, this study limited analysis to people of European genetic ancestry in the CLSA. Base GWAS data, when composed of a homogenous population (ie. European ancestry), yields higher prediction accuracy in target populations of the same (or near-same) ancestry-derived population (Wang et al., 2020) when designing polygenic risk scores. This limits the extent to which these findings can be applied to younger or more ancestrally diverse cohorts. However, it may be such that, across ages and ancestries, metabolic HPA-axis-mediated immunometabolic dysregulation causes shared disease-processes due to the common biology within the sexes. The design and application of genetic (SNP) contributions designed from one ancestry remains limited in applications on other ancestries that may have differing frequency and penetrance of those SNPs. However, if future studies analyze MDD by sex and investigate cardiometabolic risk factors as covariates, more robust inferences on any shared characteristics within-sex between ancestries and/or within-sex across age groups may be uncovered and replicated.

Additionally, diverse sex and gender information should be considered in future investigations of MDD. For example, individuals with chromosomal sexes other than 46,XX or 46,XY, and/or individuals for whom their identified gender does not match their sex assigned at birth, may have differing socioeconomic correlates and/or biological considerations. This is especially true if these individuals are taking hormone therapies that may impact their neurological and cardiovascular risk. In discussing the pathophysiology of MDD, social (gendered) causes and resulting behaviours in response to stressors has been proven to show strong influences on MDD risk and prevalence estimates (Shi et al., 2021). Socioeconomic factors such as age, partnered status, access to a stable social network and intimate social support (ie. with a partner, family, close friends), income, and education/vocation all have well-established epidemiological associations with risk of depression and IHD (Connelly et al., 2020). However, dissecting the causes and effects of these gendered risk factors was beyond the scope of this study.

## Data Availability

All data produced in the present study are available upon reasonable request to the authors.

## Acknowledgment and Disclaimers

This research was made possible using the data/biospecimens collected by the Canadian Longitudinal Study on Aging (CLSA). Funding for the Canadian Longitudinal Study on Aging (CLSA) is provided by the Government of Canada through the Canadian Institutes of Health Research (CIHR) under grant reference: LSA 94473 and the Canada Foundation for Innovation, as well as the following provinces, Newfoundland, Nova Scotia, Quebec, Ontario, Manitoba, Alberta, and British Columbia. This research has been conducted using the CLSA datasets “Baseline Tracking Dataset version 3.6” and “Baseline Comprehensive Dataset version 5.0”, under Application Number 2010011. The CLSA is led by Drs. Parminder Raina, Christina Wolfson and Susan Kirkland.

The opinions expressed in this manuscript are the authors’ own and do not reflect the views of the Canadian Longitudinal Study on Aging. The authors sincerely thank Dr. Wendy Robinson and Amy Inkster for providing helpful feedback on this manuscript.

This research was supported funding application F19-04692, sponsored by the University of British Columbia (UBC) Department of Medical Genetics with UBC Clinical Research Ethics board approval under application #H21-00050.

---

**Supplementary Table 1:** Odds ratios for MDD of participants in each decile relative to the lowest decile (referent) for each combination of input CLSA target data (females vs males) with base data (both-sexes GWAS or sex-specific GWAS).

| Sex of target CLSA data | Sex of base GWAS data | Chromosome | Quantile | Odds Ratio | 95% CI (low) | 95% CI (high) | P-value |
| --- | --- | --- | --- | --- | --- | --- | --- |
| Females | Both-sexes | Autosomes | (0-10] | REF | REF | REF | REF |
|  |  |  | (10-20] | 1.164 | 0.910 | 1.489 | 0.23 |
|  |  |  | (20-30] | 1.128 | 0.882 | 1.443 | 0.34 |
|  |  |  | (30-40] | 1.121 | 0.875 | 1.437 | 0.37 |
|  |  |  | (40-50] | 1.133 | 0.884 | 1.453 | 0.33 |
|  |  |  | (50-60] | 1.242 | 0.973 | 1.588 | 0.08 |
|  |  |  | (60-70] | 1.188 | 0.928 | 1.521 | 0.17 |
|  |  |  | (70-80] | 1.286 | 1.007 | 1.643 | 0.044 |
|  |  |  | (80-90] | 1.247 | 0.978 | 1.594 | 0.076 |
|  |  |  | (90-99] | 1.441 | 1.132 | 1.837 | 0.0031 |
| Females | Females | Autosomes | (0-10] | REF | REF | REF | REF |
|  |  |  | (10-20] | 1.339 | 1.047 | 1.714 | 0.020 |
|  |  |  | (20-30] | 1.268 | 0.989 | 1.629 | 0.062 |
|  |  |  | (30-40] | 1.255 | 0.978 | 1.613 | 0.075 |
|  |  |  | (40-50] | 1.188 | 0.922 | 1.531 | 0.18 |
|  |  |  | (50-60] | 1.300 | 1.015 | 1.668 | 0.038 |
|  |  |  | (60-70] | 1.290 | 1.006 | 1.656 | 0.045 |
|  |  |  | (70-80] | 1.419 | 1.109 | 1.818 | 0.005 |
|  |  |  | (80-90] | 1.334 | 1.040 | 1.715 | 0.024 |
|  |  |  | (90-99] | 1.676 | 1.318 | 2.136 | 0.000028 |
| Females | Males | Autosomes | (0-10] | REF | REF | REF | REF |
|  |  |  | (10-20] | 1.109 | 0.870 | 1.415 | 0.40 |
|  |  |  | (20-30] | 1.080 | 0.845 | 1.381 | 0.54 |
|  |  |  | (30-40] | 1.084 | 0.849 | 1.386 | 0.52 |
|  |  |  | (40-50] | 1.164 | 0.914 | 1.484 | 0.22 |
|  |  |  | (50-60] | 0.950 | 0.741 | 1.217 | 0.68 |
|  |  |  | (60-70] | 1.052 | 0.822 | 1.348 | 0.69 |
|  |  |  | (70-80] | 1.162 | 0.911 | 1.483 | 0.23 |
|  |  |  | (80-90] | 1.169 | 0.920 | 1.487 | 0.20 |
|  |  |  | (90-99] | 1.165 | 0.915 | 1.485 | 0.21 |
| Females | Females | X | (0-10] | REF | REF | REF | REF |
|  |  |  | (10-20] | 0.887 | 0.697 | 1.129 | 0.33 |
|  |  |  | (20-30] | 1.004 | 0.794 | 1.269 | 0.98 |
|  |  |  | (30-40] | 0.949 | 0.746 | 1.207 | 0.67 |
|  |  |  | (40-50] | 0.912 | 0.718 | 1.159 | 0.45 |
|  |  |  | (50-60] | 0.830 | 0.649 | 1.060 | 0.14 |
|  |  |  | (60-70] | 0.871 | 0.685 | 1.108 | 0.26 |
|  |  |  | (70-80] | 1.088 | 0.862 | 1.373 | 0.48 |
|  |  |  | (80-90] | 1.176 | 0.935 | 1.480 | 0.17 |
|  |  |  | (90-99] | 0.933 | 0.734 | 1.185 | 0.57 |
| Males | Both-sexes | Autosomes | (0-10] | REF | REF | REF | REF |
|  |  |  | (10-20] | 1.455 | 1.056 | 2.016 | 0.023 |
|  |  |  | (20-30] | 1.523 | 1.107 | 2.105 | 0.010 |
|  |  |  | (30-40] | 1.470 | 1.067 | 2.035 | 0.019 |
|  |  |  | (40-50] | 1.372 | 0.991 | 1.906 | 0.058 |
|  |  |  | (50-60] | 1.609 | 1.173 | 2.218 | 0.0034 |
|  |  |  | (60-70] | 1.470 | 1.067 | 2.035 | 0.019 |
|  |  |  | (70-80] | 1.506 | 1.094 | 2.083 | 0.012 |
|  |  |  | (80-90] | 1.909 | 1.404 | 2.614 | 0.000044 |
|  |  |  | (90-99] | 1.521 | 1.106 | 2.102 | 0.010 |
| Males | Females | Autosomes | (0-10] | REF | REF | REF | REF |
|  |  |  | (10-20] | 0.934 | 0.691 | 1.260 | 0.653 |
|  |  |  | (20-30] | 0.967 | 0.718 | 1.303 | 0.827 |
|  |  |  | (30-40] | 1.092 | 0.817 | 1.461 | 0.553 |
|  |  |  | (40-50] | 0.911 | 0.674 | 1.231 | 0.545 |
|  |  |  | (50-60] | 0.856 | 0.630 | 1.161 | 0.317 |
|  |  |  | (60-70] | 1.000 | 0.744 | 1.344 | 1.000 |
|  |  |  | (70-80] | 0.990 | 0.736 | 1.332 | 0.947 |
|  |  |  | (80-90] | 1.128 | 0.845 | 1.508 | 0.413 |
|  |  |  | (90-99] | 1.080 | 0.807 | 1.447 | 0.60 |
| Males | Males | Autosomes | (0-10] | REF | REF | REF | REF |
|  |  |  | (10-20] | 1.227 | 0.906 | 1.666 | 0.187 |
|  |  |  | (20-30] | 0.950 | 0.690 | 1.307 | 0.751 |
|  |  |  | (30-40] | 1.105 | 0.811 | 1.507 | 0.528 |
|  |  |  | (40-50] | 1.120 | 0.822 | 1.527 | 0.473 |
|  |  |  | (50-60] | 1.296 | 0.960 | 1.754 | 0.0918 |
|  |  |  | (60-70] | 1.158 | 0.852 | 1.576 | 0.349 |
|  |  |  | (70-80] | 1.106 | 0.812 | 1.509 | 0.522 |
|  |  |  | (80-90] | 1.421 | 1.058 | 1.916 | 0.020 |
|  |  |  | (90-99] | 1.434 | 1.068 | 1.932 | 0.017 |
| Males | Males | X | (0-10] | REF | REF | REF | REF |
|  |  |  | (10-20] | 0.990 | 0.737 | 1.330 | 0.947 |
|  |  |  | (20-30] | 1.210 | 0.911 | 1.610 | 0.190 |
|  |  |  | (30-40] | 1.011 | 0.754 | 1.357 | 0.940 |
|  |  |  | (40-50] | 1.093 | 0.818 | 1.461 | 0.549 |
|  |  |  | (50-60] | 0.945 | 0.702 | 1.273 | 0.711 |
|  |  |  | (60-70] | 0.978 | 0.727 | 1.314 | 0.880 |
|  |  |  | (70-80] | 1.070 | 0.800 | 1.431 | 0.650 |
|  |  |  | (80-90] | 0.803 | 0.589 | 1.092 | 0.163 |
|  |  |  | (90-99] | 0.760 | 0.556 | 1.037 | 0.085 |

**Supplementary Table 2:** Summary of the number of missing data in each of the finalized covariates prior to sample QC filtering out samples with missing values in any covariates.

| Variable | Labels (if categorical) | Original labels in CLSA (prior to recoding) | Females (n=13, 221) |  | Males (n=13, 280) |  |
| --- | --- | --- | --- | --- | --- | --- |
|  |  |  | Count of NAs | Total (%) | Count of NAs | Total (%) |
| MDD | Don't Know/No answer | 8 | 43 | 58 (0.4) | 33 | 63 (0.5) |
|  | Refused | 9 | 2 |  | 3 |  |
|  | NA | NA | 13 |  | 27 |  |
| Sex |  |  | 0 | 0 | 0 | 0 |
| Age |  | # | 0 | 0 | 0 | 0 |
| Partnered Status | Don't Know/No answer | 8 | 2 | 693 (5.2) | 3 | 762 (5.7) |
|  | Refused | 9 | 1 |  | 3 |  |
|  | Missing - Technical difficulties | -8 | 168 |  | 206 |  |
|  | NA | NA | 522 |  | 550 |  |
| Annual Income | Don't Know/No answer | 8 | 471 | 1027 (7.8) | 214 | 623 (4.7) |
|  | Refused | 9 | 556 |  | 409 |  |
|  | Missing - Technical difficulties | 7 | 0 |  | 0 |  |
| Highest Education | Other | 97 | 17 | 2199 (16.6) | 16 | 1786 (13.4) |
|  | Don't Know/No answer | 98 | 1 |  | 2 |  |
|  | Refused | 99 | 0 |  | 0 |  |
|  | NA | NA | 2181 |  | 1768 |  |
| Smoking Status | Missing | -8 | 0 | 0 | 0 | 0 |
| Frequency of alcohol consumption | Don't Know/No answer | 98 | 4 | 390 (2.9) | 4 | 233 (1.8) |
|  | Refused | 99 | 1 |  | 1 |  |
|  | Missing | 77 | 0 |  | 0 |  |
|  | NA | NA | 385 |  | 228 |  |
| High Blood Pressure | Don't Know/No answer | 8 | 34 | 34 (0.3) | 63 | 63 (0.5) |
|  | Refused | 9 | 0 |  | 0 |  |
| Diabetes | Don't Know/No answer | 8 | 18 | 18 (0.1) | 24 | 24 (0.2) |
|  | Refused | 9 | 0 |  | 0 |  |
| Hypothyroidism | Don't Know/No answer | 8 | 160 | 160 (1.2) | 118 | 118 (0.9) |
|  | Refused | 9 | 0 |  | 0 |  |
| Waist-to-hip ratio |  | 999.7 | 27 | 27 (0.2) | 27 | 43 (0.3) |
| Angina | Don't Know/No answer | 8 | 45 | 45 (0.3) | 44 | 44 (0.3) |
|  | Refused | 9 | 0 |  | 0 |  |
| MI | Don't Know/No answer | 8 | 19 | 19 (0.1) | 44 | 44 (0.3) |
|  | Refused | 9 | 0 |  | 0 |  |

**Supplementary Table 3:** Correlation matrix of covariates for females.

|  | Age | Partner<br>ed<br>Status | Annual<br>Income | Highest<br>Educati<br>on | Smokin<br>g Status | Alcohol<br>Frequen<br>cy | High<br>Blood<br>Pressur<br>e | Diabete<br>s | Hypoth<br>yroidis<br>m | WHR | LLT | IHD | MDD |
| --- | --- | --- | --- | --- | --- | --- | --- | --- | --- | --- | --- | --- | --- |
| Age | 1 | 0.24 | 0.36 | 0.13 | -0.01 | -0.06 | 0.27 | 0.08 | 0.11 | 0.16 | 0.22 | 0.16 | -0.09 |
| Partner<br>ed<br>Status | 0.24 | 1 | 0.51 | 0.06 | 0.09 | -0.05 | 0.09 | 0.06 | 0.06 | 0.1 | 0.09 | 0.06 | 0.1 |
| Annual<br>Income | 0.36 | 0.51 | 1 | 0.25 | 0.1 | -0.13 | 0.15 | 0.11 | 0.08 | 0.14 | 0.14 | 0.1 | 0.11 |
| Highest<br>Educati<br>on | 0.13 | 0.06 | 0.25 | 1 | 0.15 | -0.06 | 0.09 | 0.09 | 0.02 | 0.11 | 0.08 | 0.06 | 0.01 |
| Smokin<br>g Status | -0.01 | 0.09 | 0.1 | 0.15 | 1 | 0.01 | 0.02 | 0.03 | -0.02 | 0.08 | 0.03 | 0.05 | 0.07 |
| Alcohol<br>Frequen<br>cy | -0.06 | -0.05 | -0.13 | -0.06 | 0.01 | 1 | -0.04 | -0.08 | 0 | -0.02 | -0.05 | -0.05 | -0.05 |
| High<br>Blood<br>Pressur<br>e | 0.27 | 0.09 | 0.15 | 0.09 | 0.02 | -0.04 | 1 | 0.22 | 0.03 | 0.23 | 0.24 | 0.12 | 0.02 |
| Diabete<br>s | 0.08 | 0.06 | 0.11 | 0.09 | 0.03 | -0.08 | 0.22 | 1 | 0.04 | 0.23 | 0.22 | 0.07 | 0.07 |
| Hypoth<br>yroidis<br>m | 0.11 | 0.06 | 0.08 | 0.02 | -0.02 | 0 | 0.03 | 0.04 | 1 | 0.06 | 0.05 | 0.03 | 0.06 |
| WHR | 0.16 | 0.1 | 0.14 | 0.11 | 0.08 | -0.02 | 0.23 | 0.23 | 0.06 | 1 | 0.19 | 0.08 | 0.06 |
| LLT | 0.22 | 0.09 | 0.14 | 0.08 | 0.03 | -0.05 | 0.24 | 0.22 | 0.05 | 0.19 | 1 | 0.23 | 0.04 |
| IHD | 0.16 | 0.06 | 0.1 | 0.06 | 0.05 | -0.05 | 0.12 | 0.07 | 0.03 | 0.08 | 0.23 | 1 | 0.04 |
| MDD | -0.09 | 0.1 | 0.11 | 0.01 | 0.07 | -0.05 | 0.02 | 0.07 | 0.06 | 0.06 | 0.04 | 0.04 | 1 |

**Supplementary Table 4:** P-values of correlation matrix of covariates for females.

|  | Age | Partner<br>ed<br>Status | Annual<br>Income | Highest<br>Educati<br>on | Smokin<br>g Status | Alcohol<br>Frequen<br>cy | High<br>Blood<br>Pressur<br>e | Diabete<br>s | Hypoth<br>yroidis<br>m | WHR | LLT | IHD | MDD |
| --- | --- | --- | --- | --- | --- | --- | --- | --- | --- | --- | --- | --- | --- |
| Age | 0.00 | 0.33 | 0.11 | 0.85 | 0.38 | 0.29 | 0.29 | 0.88 | 0.99 | 0.79 | 0.47 | 0.72 | 0.18 |
| Partner<br>ed<br>Status | 0.33 | 0.00 | 0.01 | 0.89 | 0.90 | 0.34 | 0.84 | 0.68 | 0.79 | 0.83 | 0.79 | 0.73 | 1.00 |
| Annual<br>Income | 0.11 | 0.01 | 0.00 | 0.43 | 0.86 | 0.11 | 0.91 | 0.85 | 0.79 | 0.97 | 0.99 | 0.86 | 0.90 |
| Highest<br>Educati<br>on | 0.85 | 0.89 | 0.43 | 0.00 | 0.70 | 0.38 | 0.88 | 0.91 | 0.60 | 0.96 | 0.80 | 0.78 | 0.59 |
| Smokin<br>g Status | 0.38 | 0.90 | 0.86 | 0.70 | 0.00 | 0.81 | 0.47 | 0.60 | 0.45 | 0.76 | 0.50 | 0.71 | 0.98 |
| Alcohol<br>Frequen<br>cy | 0.29 | 0.34 | 0.11 | 0.38 | 0.81 | 0.00 | 0.38 | 0.31 | 0.77 | 0.46 | 0.34 | 0.44 | 0.56 |
| High<br>Blood<br>Pressur<br>e | 0.29 | 0.84 | 0.91 | 0.88 | 0.47 | 0.38 | 0.00 | 0.41 | 0.60 | 0.41 | 0.34 | 0.89 | 0.49 |
| Diabete<br>s | 0.88 | 0.68 | 0.85 | 0.91 | 0.60 | 0.31 | 0.41 | 0.00 | 0.67 | 0.38 | 0.43 | 0.87 | 0.86 |
| Hypoth<br>yroidis<br>m | 0.99 | 0.79 | 0.79 | 0.60 | 0.45 | 0.77 | 0.60 | 0.67 | 0.00 | 0.70 | 0.66 | 0.66 | 0.90 |
| WHR | 0.79 | 0.83 | 0.97 | 0.96 | 0.76 | 0.46 | 0.41 | 0.38 | 0.70 | 0.00 | 0.61 | 0.83 | 0.71 |
| LLT | 0.47 | 0.79 | 0.99 | 0.80 | 0.50 | 0.34 | 0.34 | 0.43 | 0.66 | 0.61 | 0.00 | 0.39 | 0.59 |
| IHD | 0.72 | 0.73 | 0.86 | 0.78 | 0.71 | 0.44 | 0.89 | 0.87 | 0.66 | 0.83 | 0.39 | 0.00 | 0.70 |
| MDD | 0.18 | 1.00 | 0.90 | 0.59 | 0.98 | 0.56 | 0.49 | 0.86 | 0.90 | 0.71 | 0.59 | 0.70 | 0.00 |

**Supplementary Table 5:** Correlation matrix of covariates for males.

|  | Age | Partner<br>ed<br>Status | Annual<br>Income | Highest<br>Educati<br>on | Smokin<br>g Status | Alcohol<br>Frequen<br>cy | High<br>Blood<br>Pressur<br>e | Diabete<br>s | Hypoth<br>yroidis<br>m | WHR | LLT | IHD | MDD |
| --- | --- | --- | --- | --- | --- | --- | --- | --- | --- | --- | --- | --- | --- |
| Age | 1 | 0.04 | 0.27 | 0.01 | 0.1 | -0.04 | 0.23 | 0.13 | 0.1 | 0.08 | 0.26 | 0.2 | -0.06 |
| Partner<br>ed<br>Status | 0.04 | 1 | 0.39 | 0.07 | 0.08 | -0.06 | 0.02 | 0.03 | 0.02 | 0.04 | -0.01 | 0.02 | 0.07 |
| Annual<br>Income | 0.27 | 0.39 | 1 | 0.23 | 0.14 | -0.14 | 0.09 | 0.11 | 0.05 | 0.07 | 0.09 | 0.09 | 0.09 |
| Highest<br>Educati<br>on | 0.01 | 0.07 | 0.23 | 1 | 0.17 | -0.07 | 0.05 | 0.09 | 0.01 | 0.1 | 0.05 | 0.05 | 0.01 |
| Smokin<br>g Status | 0.1 | 0.08 | 0.14 | 0.17 | 1 | -0.01 | 0.08 | 0.08 | 0 | 0.12 | 0.07 | 0.06 | 0.06 |
| Alcohol<br>Frequen<br>cy | -0.04 | -0.06 | -0.14 | -0.07 | -0.01 | 1 | -0.01 | -0.05 | -0.01 | -0.04 | -0.01 | -0.02 | -0.08 |
| High<br>Blood<br>Pressur<br>e | 0.23 | 0.02 | 0.09 | 0.05 | 0.08 | -0.01 | 1 | 0.21 | 0.05 | 0.19 | 0.25 | 0.14 | 0.05 |
| Diabete<br>s | 0.13 | 0.03 | 0.11 | 0.09 | 0.08 | -0.05 | 0.21 | 1 | 0.03 | 0.18 | 0.25 | 0.12 | 0.05 |
| Hypothyroidism | 0.1 | 0.02 | 0.05 | 0.01 | 0 | -0.01 | 0.05 | 0.03 | 1 | 0.02 | 0.07 | 0.04 | 0.02 |
| WHR | 0.08 | 0.04 | 0.07 | 0.1 | 0.12 | -0.04 | 0.19 | 0.18 | 0.02 | 1 | 0.12 | 0.08 | 0.07 |
| LLT | 0.26 | -0.01 | 0.09 | 0.05 | 0.07 | -0.01 | 0.25 | 0.25 | 0.07 | 0.12 | 1 | 0.3 | 0.01 |
| IHD | 0.2 | 0.02 | 0.09 | 0.05 | 0.06 | -0.02 | 0.14 | 0.12 | 0.04 | 0.08 | 0.3 | 1 | 0.03 |
| MDD | -0.06 | 0.07 | 0.09 | 0.01 | 0.06 | -0.08 | 0.05 | 0.05 | 0.02 | 0.07 | 0.01 | 0.03 | 1 |

**Supplementary Table 6:**
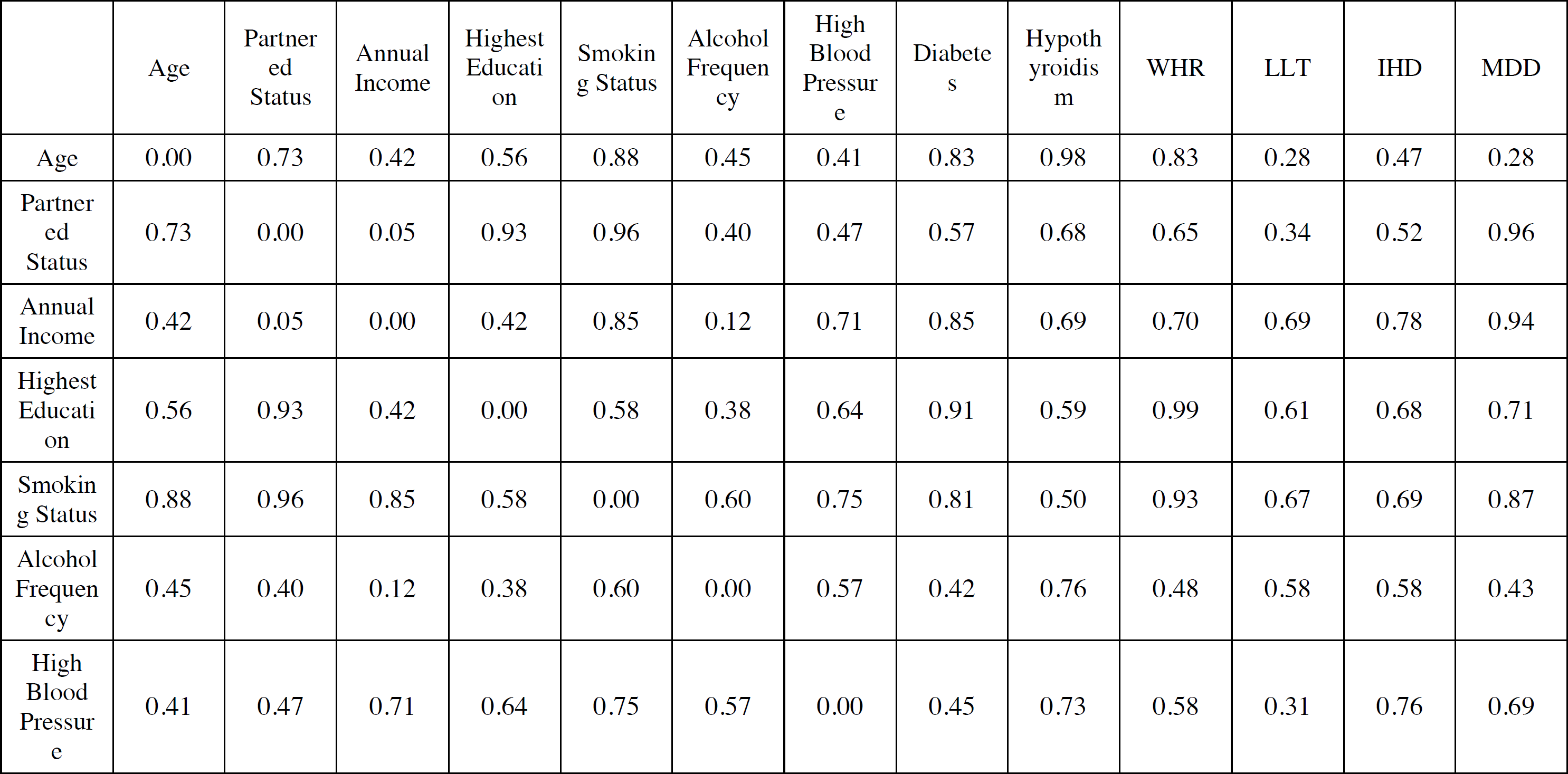
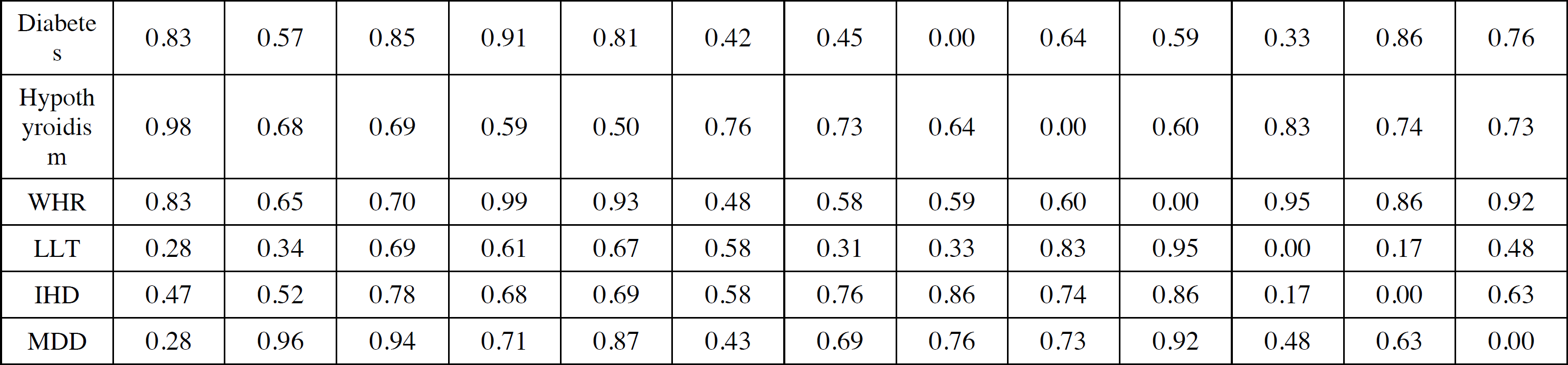
P-values of correlation matrix of covariates for males.

**Supplementary Table 7:**
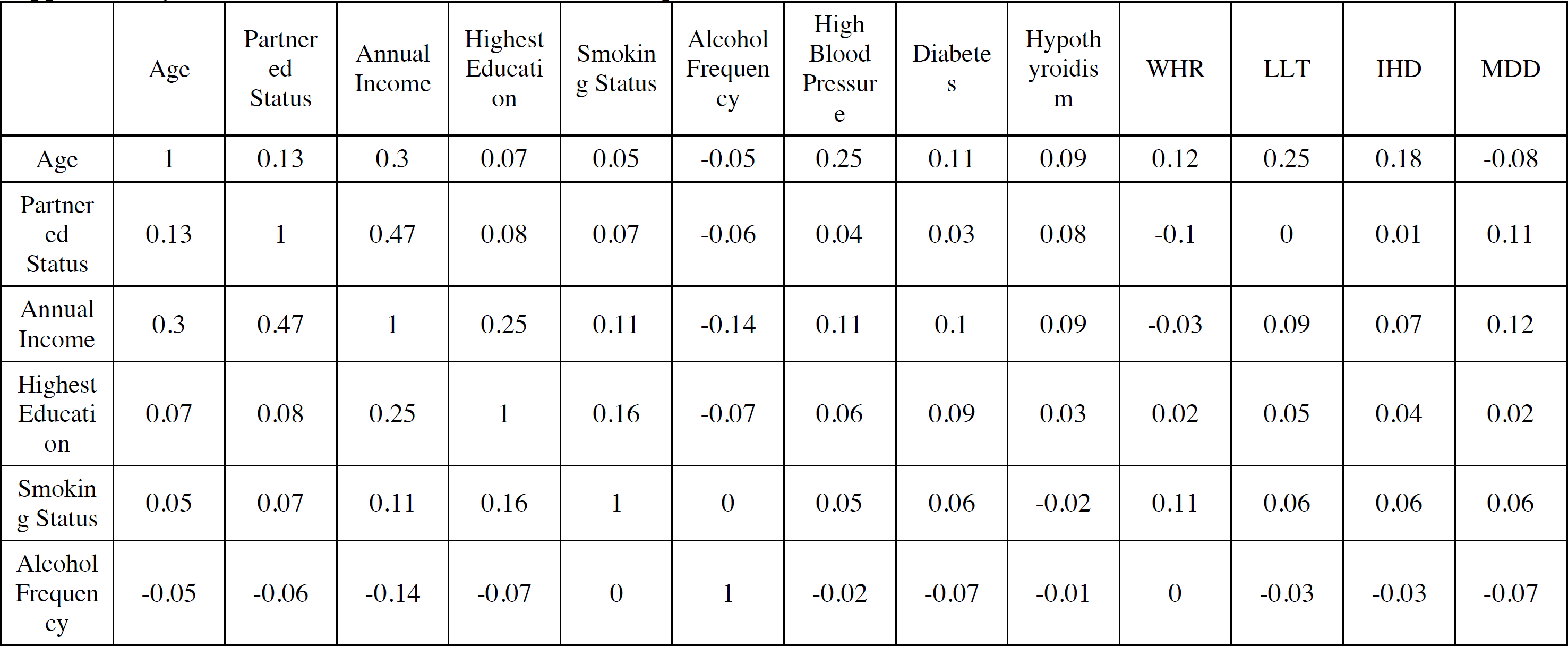
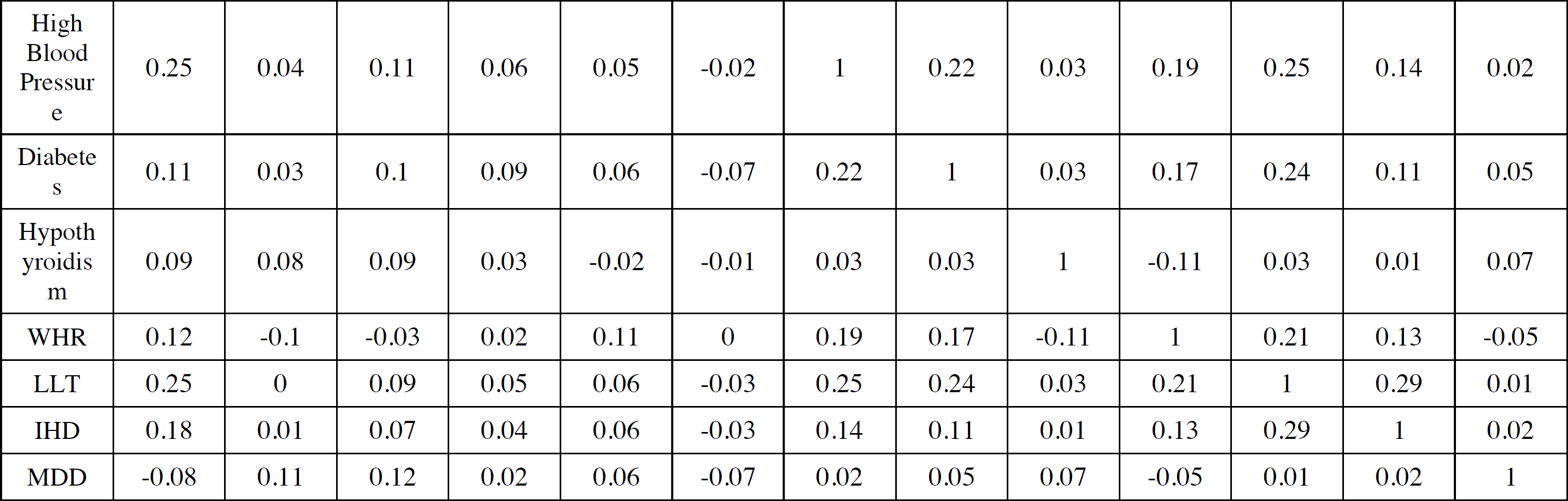
Correlation matrix of covariates of pooled “both-sexes” data.

|  | Age | Partner<br>ed<br>Status | Annual<br>Income | Highest<br>Educati<br>on | Smokin<br>g Status | Alcohol<br>Frequen<br>cy | High<br>Blood<br>Pressur<br>e | Diabetes | Hypoth<br>yroidis<br>m | WHR | LLT | IHD | MDD |
| --- | --- | --- | --- | --- | --- | --- | --- | --- | --- | --- | --- | --- | --- |
| Age | 1 | 0.13 | 0.3 | 0.07 | 0.05 | -0.05 | 0.25 | 0.11 | 0.09 | 0.12 | 0.25 | 0.18 | -0.08 |
| Partner<br>ed<br>Status | 0.13 | 1 | 0.47 | 0.08 | 0.07 | -0.06 | 0.04 | 0.03 | 0.08 | -0.1 | 0 | 0.01 | 0.11 |
| Annual<br>Income | 0.3 | 0.47 | 1 | 0.25 | 0.11 | -0.14 | 0.11 | 0.1 | 0.09 | -0.03 | 0.09 | 0.07 | 0.12 |
| Highest<br>Educati<br>on | 0.07 | 0.08 | 0.25 | 1 | 0.16 | -0.07 | 0.06 | 0.09 | 0.03 | 0.02 | 0.05 | 0.04 | 0.02 |
| Smokin<br>g Status | 0.05 | 0.07 | 0.11 | 0.16 | 1 | 0 | 0.05 | 0.06 | -0.02 | 0.11 | 0.06 | 0.06 | 0.06 |
| Alcohol<br>Frequen<br>cy | -0.05 | -0.06 | -0.14 | -0.07 | 0 | 1 | -0.02 | -0.07 | -0.01 | 0 | -0.03 | -0.03 | -0.07 |
| High Blood Pressure | 0.25 | 0.04 | 0.11 | 0.06 | 0.05 | -0.02 | 1 | 0.22 | 0.03 | 0.19 | 0.25 | 0.14 | 0.02 |
| Diabetes | 0.11 | 0.03 | 0.1 | 0.09 | 0.06 | -0.07 | 0.22 | 1 | 0.03 | 0.17 | 0.24 | 0.11 | 0.05 |
| Hypothyroidism | 0.09 | 0.08 | 0.09 | 0.03 | -0.02 | -0.01 | 0.03 | 0.03 | 1 | -0.11 | 0.03 | 0.01 | 0.07 |
| WHR | 0.12 | -0.1 | -0.03 | 0.02 | 0.11 | 0 | 0.19 | 0.17 | -0.11 | 1 | 0.21 | 0.13 | -0.05 |
| LLT | 0.25 | 0 | 0.09 | 0.05 | 0.06 | -0.03 | 0.25 | 0.24 | 0.03 | 0.21 | 1 | 0.29 | 0.01 |
| IHD | 0.18 | 0.01 | 0.07 | 0.04 | 0.06 | -0.03 | 0.14 | 0.11 | 0.01 | 0.13 | 0.29 | 1 | 0.02 |
| MDD | -0.08 | 0.11 | 0.12 | 0.02 | 0.06 | -0.07 | 0.02 | 0.05 | 0.07 | -0.05 | 0.01 | 0.02 | 1 |

**Supplementary Table 8:** P-values of correlation matrix of pooled “both-sexes” data.

|  | Age | Partnered Status | Annual Income | Highest Education | Smoking Status | Alcohol Frequency | High Blood Pressure | Diabetes | Hypothyroidism | WHR | LLT | IHD | MDD |
| --- | --- | --- | --- | --- | --- | --- | --- | --- | --- | --- | --- | --- | --- |
| Age | 0.00 | 0.81 | 0.30 | 0.85 | 0.63 | 0.36 | 0.34 | 0.93 | 0.93 | 0.85 | 0.34 | 0.58 | 0.23 |
| Partnered Status | 0.81 | 0.00 | 0.01 | 0.86 | 0.88 | 0.38 | 0.55 | 0.55 | 0.87 | 0.14 | 0.35 | 0.47 | 0.72 |
| Annual Income | 0.30 | 0.01 | 0.00 | 0.35 | 0.96 | 0.12 | 0.81 | 0.79 | 0.96 | 0.26 | 0.64 | 0.65 | 0.85 |
| Highest Education | 0.85 | 0.86 | 0.35 | 0.00 | 0.62 | 0.37 | 0.69 | 0.90 | 0.76 | 0.62 | 0.61 | 0.65 | 0.77 |
| Smoking Status | 0.63 | 0.88 | 0.96 | 0.62 | 0.00 | 0.71 | 0.63 | 0.74 | 0.46 | 0.91 | 0.66 | 0.74 | 0.90 |
| Alcohol<br>Frequen<br>cy | 0.36 | 0.38 | 0.12 | 0.37 | 0.71 | 0.00 | 0.51 | 0.35 | 0.74 | 0.80 | 0.48 | 0.55 | 0.48 |
| High<br>Blood<br>Pressur<br>e | 0.34 | 0.55 | 0.81 | 0.69 | 0.63 | 0.51 | 0.00 | 0.40 | 0.59 | 0.43 | 0.28 | 0.71 | 0.51 |
| Diabete<br>s | 0.93 | 0.55 | 0.79 | 0.90 | 0.74 | 0.35 | 0.40 | 0.00 | 0.62 | 0.51 | 0.34 | 0.86 | 0.74 |
| Hypoth<br>yroidis<br>m | 0.93 | 0.87 | 0.96 | 0.76 | 0.46 | 0.74 | 0.59 | 0.62 | 0.00 | 0.18 | 0.55 | 0.56 | 0.91 |
| WHR | 0.85 | 0.14 | 0.26 | 0.62 | 0.91 | 0.80 | 0.43 | 0.51 | 0.18 | 0.00 | 0.34 | 0.64 | 0.34 |
| LLT | 0.34 | 0.35 | 0.64 | 0.61 | 0.66 | 0.48 | 0.28 | 0.34 | 0.55 | 0.34 | 0.00 | 0.18 | 0.44 |
| IHD | 0.58 | 0.47 | 0.65 | 0.65 | 0.74 | 0.55 | 0.71 | 0.86 | 0.56 | 0.64 | 0.18 | 0.00 | 0.58 |
| MDD | 0.23 | 0.72 | 0.85 | 0.77 | 0.90 | 0.48 | 0.51 | 0.74 | 0.91 | 0.34 | 0.44 | 0.58 | 0.00 |

**Supplementary Table 9:**
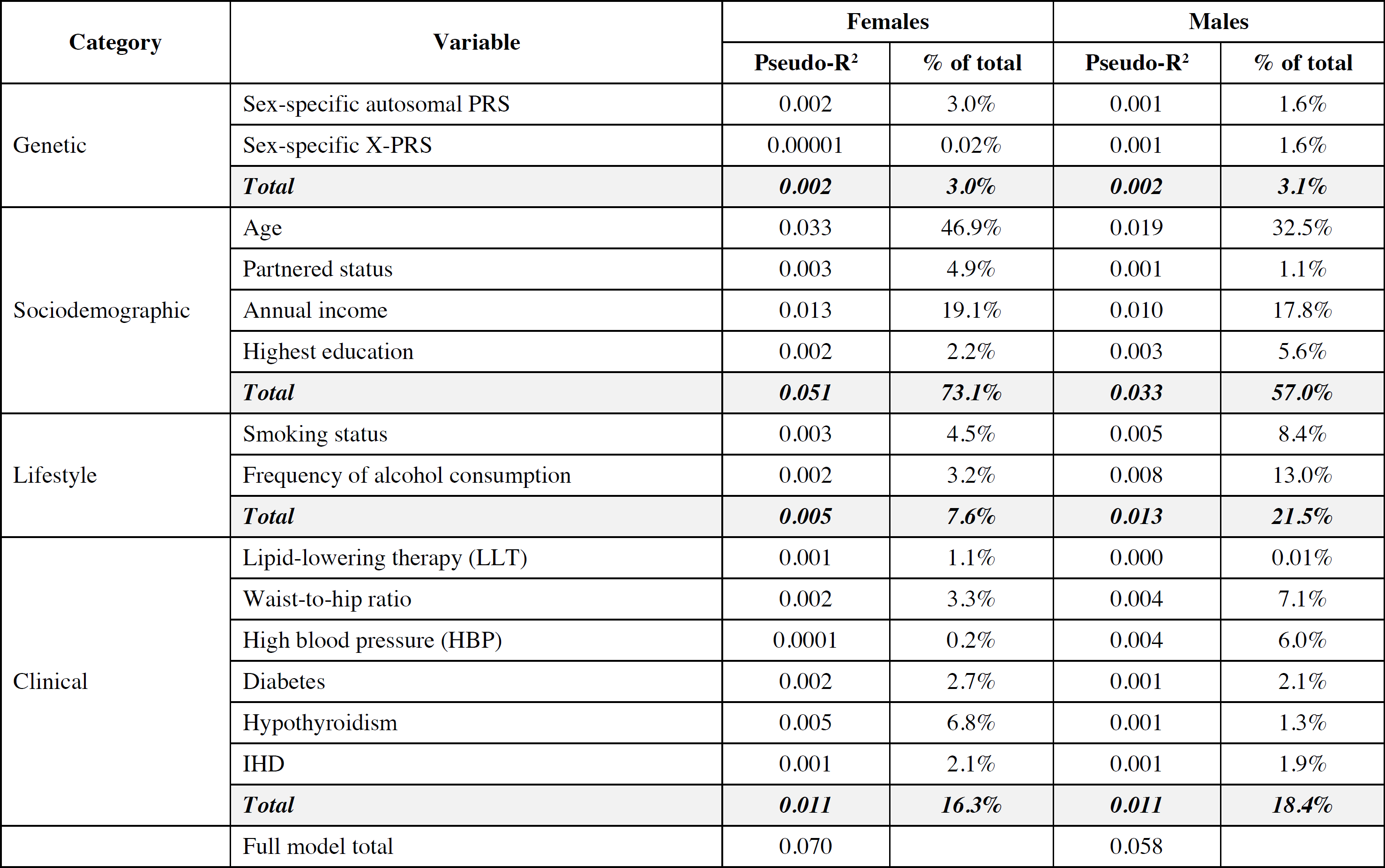
Pseudo-R^2^ for each variable reported as a percent (%) of MDD explained, as well as a relative % to the other categories to the total.

| Category | Variable | Females |  | Males |  |
| --- | --- | --- | --- | --- | --- |
|  |  | Pseudo-R<sup>2</sup> | % of total | Pseudo-R<sup>2</sup> | % of total |
| Genetic | Sex-specific autosomal PRS | 0.002 | 3.0% | 0.001 | 1.6% |
|  | Sex-specific X-PRS | 0.00001 | 0.02% | 0.001 | 1.6% |
|  | **Total** | **0.002** | **3.0%** | **0.002** | **3.1%** |
| Sociodemographic | Age | 0.033 | 46.9% | 0.019 | 32.5% |
|  | Partnered status | 0.003 | 4.9% | 0.001 | 1.1% |
|  | Annual income | 0.013 | 19.1% | 0.010 | 17.8% |
|  | Highest education | 0.002 | 2.2% | 0.003 | 5.6% |
|  | **Total** | **0.051** | **73.1%** | **0.033** | **57.0%** |
| Lifestyle | Smoking status | 0.003 | 4.5% | 0.005 | 8.4% |
|  | Frequency of alcohol consumption | 0.002 | 3.2% | 0.008 | 13.0% |
|  | **Total** | **0.005** | **7.6%** | **0.013** | **21.5%** |
| Clinical | Lipid-lowering therapy (LLT) | 0.001 | 1.1% | 0.000 | 0.01% |
|  | Waist-to-hip ratio | 0.002 | 3.3% | 0.004 | 7.1% |
|  | High blood pressure (HBP) | 0.0001 | 0.2% | 0.004 | 6.0% |
|  | Diabetes | 0.002 | 2.7% | 0.001 | 2.1% |
|  | Hypothyroidism | 0.005 | 6.8% | 0.001 | 1.3% |
|  | IHD | 0.001 | 2.1% | 0.001 | 1.9% |
|  | **Total** | **0.011** | **16.3%** | **0.011** | **18.4%** |
|  | Full model total | 0.070 |  | 0.058 |  |

